# Efficacy and Safety of IL-23 Inhibitors in the Treatment of Crohn’s Disease and Ulcerative Colitis: Systemic Review and Network Meta-Analysis

**DOI:** 10.1101/2025.09.03.25334996

**Authors:** Ritik Kaste, Richa Thyagarajan, Chidurala Rahul, Gaurav Sharma, G Saicharan Ashrith, Mohammed Sahal, Deep Patel, Vrund Patel, Ashesh Das, Harshwardhan Dhanraj Ramteke, Rakhshanda Khan

## Abstract

**Introduction:** Inflammatory Bowel Disease (IBD), which includes Crohn’s Disease (CD) and Ulcerative Colitis (UC), is a chronic condition that causes inflammation in the gastrointestinal tract and has been steadily increasing in prevalence globally. Traditional therapies for IBD often have limitations in efficacy and long-term safety. In recent years, interleukin-23 (IL-23) inhibitors, such as Ustekinumab, Risankizumab, Guselkumab, and Mirikizumab, have shown promise in treating IBD by targeting immune pathways involved in disease progression. This network meta-analysis (NMA) aims to evaluate the efficacy and safety of these IL-23 inhibitors in treating both Crohn’s Disease and Ulcerative Colitis.

**Methods:** A comprehensive literature search was conducted across PubMed, Scopus, and the Cochrane Library to identify randomized controlled trials (RCTs) and observational studies published from 2000 to 2023 that assessed the efficacy and safety of IL-23 inhibitors in treating CD and UC. The studies selected focused on clinical outcomes such as remission rates, mucosal healing, and adverse events, comparing Ustekinumab, Risankizumab, Guselkumab, and Mirikizumab to placebo. Data were extracted for adult patients diagnosed with CD and UC, evaluating the results from the induction and maintenance phases. Statistical analysis was conducted using a random-effects model to account for heterogeneity, and I² statistics were used to assess variability across the studies.

**Results:** A total of 33 studies were included in the analysis, encompassing 19,668 patients, with a male predominance (53.6%). The efficacy of IL-23 inhibitors was compared across both CD and UC induction and maintenance phases. For CD induction, Mirikizumab showed the highest efficacy (OR = 5.19, 95% CI: 1.75 to 16.7), followed by Guselkumab (OR = 3.07, 95% CI: 1.35 to 5.98). In UC induction, Guselkumab demonstrated the most significant benefit (OR = 3.80, 95% CI: 1.10 to 13.3), with Risankizumab closely following (OR = 3.96, 95% CI: 0.685 to 23.0). During maintenance treatment, Guselkumab showed the highest odds ratio for both CD (OR = 10.3, 95% CI: 2.46 to 45.9) and UC (OR = 3.07, 95% CI: 0.698 to 13.4). Regarding safety, Risankizumab and Mirikizumab were associated with the most significant reductions in nausea and infections, while Ustekinumab and Guselkumab showed fewer benefits in these adverse events. Mirikizumab and Risankizumab also demonstrated the most substantial improvements in Quality of Life (QOL) for patients with CD and UC, while Guselkumab had mixed results, and Ustekinumab showed limited improvements in QOL.

**Conclusion:** The results of this network meta-analysis indicate that Mirikizumab and Risankizumab are the most effective IL-23 inhibitors for achieving remission and improving QOL in Crohn’s Disease and Ulcerative Colitis, with Guselkumab also showing strong efficacy, particularly in UC. Ustekinumab demonstrated more modest effects. Both Risankizumab and Mirikizumab were associated with favorable safety profiles, significantly reducing nausea and infection rates. These findings support the use of Mirikizumab and Risankizumab as potential first-line biologic therapies for patients with moderate to severe IBD.

## Introduction

Inflammatory Bowel Disease (IBD), encompassing Crohn’s Disease (CD) and Ulcerative Colitis (UC), represents a chronic, relapsing condition characterized by widespread inflammation of the gastrointestinal tract. Over the past few decades, the global prevalence of IBD has been increasing at an alarming rate [1]. Current estimates suggest that 6–8 million people worldwide are living with IBD, with Western countries, particularly the United States and Europe, reporting the highest prevalence rates [2]. The U.S. alone is home to over 1.3 million IBD patients, while nations in Asia and the Middle East are experiencing a rise in incidence, largely due to urbanization and changing dietary patterns [3]. IBD typically manifests in young adults, with a peak onset between the ages of 15 and 40, presenting a significant challenge to public health systems globally [4]. The economic burden associated with IBD is substantial, with treatment costs continuing to escalate. In 2023, the direct and indirect costs of IBD management reached an estimated $5.38 billion in Canada, and $8.5 billion annually in the U.S., driven largely by the high cost of biologics [5]. In Japan, the average monthly treatment cost for UC patients was ¥76,374 ($700), underscoring the financial burden associated with ongoing disease management [6].

The management of IBD primarily involves reducing inflammation, inducing and maintaining remission, and preventing disease relapse. Conventional therapies, such as aminosalicylates (5-ASA), corticosteroids, and immunosuppressive agents, have been the mainstay of treatment for decades [7]. Aminosalicylates, such as mesalamine, are often used as first-line therapies in mild to moderate UC, providing good efficacy with relatively fewer side effects. However, they are less effective in patients with severe disease or CD, limiting their broader applicability. Corticosteroids, although effective in managing acute flare-ups, carry significant long-term side effects, including osteoporosis, weight gain, and increased susceptibility to infections, making them unsuitable for long-term management. Immunosuppressive agents like azathioprine and methotrexate help maintain remission but require months to show benefits and are associated with risks, such as bone marrow suppression and liver toxicity. The limitations of these conventional therapies highlight the need for more effective and targeted treatment options for IBD patients.

Recent advancements in the understanding of IBD pathogenesis have led to the development of biologic therapies that specifically target the underlying immune mechanisms driving disease progression. One such breakthrough in IBD management is the targeting of interleukin-23 (IL-23), a pro-inflammatory cytokine implicated in the pathogenesis of both CD and UC. IL-23 consists of two subunits: p40 and p19 [8]. While the p40 subunit is shared with IL-12, the p19 subunit is unique to IL-23 and plays a crucial role in the activation of Th17 cells. These Th17 cells, in turn, produce cytokines such as IL-17, IL-21, and IL-22, which are key contributors to the chronic inflammation and tissue damage observed in IBD. Elevated levels of IL-23 have been found in both the serum and intestinal tissues of IBD patients, correlating with disease activity and severity, making it an attractive target for therapeutic intervention.

Several IL-23 inhibitors have been developed and are being explored in clinical trials for IBD. Ustekinumab, which targets both IL-12 and IL-23, has been approved for the treatment of CD and UC, showing significant efficacy in inducing and maintaining remission. Risankizumab, a selective inhibitor of the p19 subunit of IL-23, has demonstrated promising results in clinical trials for CD, with improved disease outcomes and a favorable safety profile. Other IL-23 inhibitors, such as Guselkumab and Mirikizumab, are also being investigated for their potential use in IBD, showing similar promising efficacy and safety profiles [9]. These targeted therapies offer several advantages over traditional treatments, including more effective disease control, fewer side effects, and a more precise mechanism of action.

Despite the promising potential of IL-23 inhibitors, comparative effectiveness and safety data across these therapies remain limited. To address this gap, a network meta-analysis (NMA) is an invaluable tool to assess and compare the efficacy and safety of multiple IL-23 inhibitors in the treatment of CD and UC. This NMA will integrate data from randomized controlled trials (RCTs) to evaluate the relative effectiveness of IL-23 inhibitors, including ustekinumab, risankizumab, guselkumab, and mirikizumab, with respect to key clinical outcomes such as remission rates, mucosal healing, and adverse events. Moreover, this analysis will explore the safety profiles of these biologics, considering adverse events such as infections, malignancies, and other long-term risks associated with immunomodulatory therapies.

The rise in IBD prevalence, coupled with the limitations of conventional therapies, underscores the need for more effective and safer treatment options. IL-23 inhibitors, with their targeted action and promising clinical trial results, have emerged as a new frontier in IBD therapy. This network meta-analysis aims to provide a comprehensive evaluation of the efficacy and safety of IL-23 inhibitors, thereby informing clinical decision-making and contributing to the growing body of evidence on biologic therapies for IBD.

## Methods

### Literature search

A comprehensive literature search was conducted across major databases including PubMed, Scopus, and Cochrane Library to identify randomized controlled trials (RCTs) and observational studies evaluating the efficacy and safety of IL-23 inhibitors in the treatment of Crohn’s disease (CD) and ulcerative colitis (UC). Keywords such as "IL-23 inhibitors," "ustekinumab," "risankizumab," "guselkumab," "mirikizumab," "Crohn’s disease," "ulcerative colitis," and "biologic therapy" were used. Studies published in English from January 2000 to present were included. Relevant clinical outcomes such as remission rates, adverse events, and mucosal healing were extracted for analysis in this network meta-analysis. The protocol for this review was registered with PROSPERO (registration number CRD420251138575). The literature search followed the PRISMA protocols [10].

### Study Selection and Data Extraction

The study selection and data extraction process will be conducted in a systematic and rigorous manner to ensure the inclusion of relevant and high-quality studies. Eligible studies must be randomized controlled trials (RCTs) that include adult patients (18 years and older) diagnosed with Crohn’s disease (CD) or ulcerative colitis (UC), and who have received treatment with IL-23 inhibitors such as ustekinumab, risankizumab, guselkumab, or mirikizumab. Only studies that report on clinical outcomes, including remission rates, mucosal healing, clinical response, disease flare-ups, and adverse events, will be included. Studies with a minimum follow-up period of 8 weeks will be considered, and only English-language publications will be reviewed. Exclusion criteria include studies involving pediatric populations, non-human studies, and studies without relevant clinical or safety data.

Two independent reviewers will extract data from the selected studies using a standardized form, ensuring accuracy and consistency in the process. The data to be extracted includes study characteristics (e.g., study design, sample size, patient demographics), details of the intervention (e.g., IL-23 inhibitor used, dosage, treatment duration), and primary and secondary efficacy outcomes, such as remission rates, mucosal healing, clinical response, and disease flare-ups. Safety outcomes, including adverse events, serious adverse events, and long-term safety concerns, will also be extracted. Statistical data, such as effect sizes (odds ratios, risk ratios, mean differences), standard deviations, and confidence intervals, will be recorded for each outcome.

If there is missing or incomplete data, efforts will be made to contact the authors for clarification. In cases where data cannot be obtained, the study will be excluded. The risk of bias for each study will be assessed using the Cochrane Risk of Bias tool for RCTs. This assessment will ensure the reliability and validity of the data used in the network meta-analysis. By following this systematic approach, the review will provide a robust and comprehensive evaluation of the efficacy and safety of IL-23 inhibitors in IBD treatment.

### Risk of Bias

The risk of bias for each study was assessed using the Cochrane Risk of Bias tool for RCTs and ROBS 2.0 for observational studies [11]. Domains evaluated included randomization, deviations from interventions, missing data, outcome measurement, and reporting bias. Studies were rated for risk of bias and categorized as low, moderate, or high.

### Statistical Analysis

For the systematic review and network meta-analysis, a random-effects model was used to account for heterogeneity across studies. Direct and indirect comparisons were conducted to evaluate the efficacy and safety of different medical treatments for Cushing’s disease. Effect sizes, including odds ratios (ORs) for binary outcomes and mean differences (MDs) for continuous outcomes, were calculated. To assess statistical heterogeneity, I² statistics were used, with values greater than 50% indicating substantial heterogeneity. Sensitivity analyses were performed to test the robustness of the findings, and publication bias was assessed using funnel plots and Egger’s test. All analyses were conducted using the R software package (version 4.0.3) and the ‘gemtc’ package for network meta-analysis.

## Results

### Demographics

A total of 2512 studies were analyzed out of which 33 studies were selected [12-29] (Figure 1). The total population across the included studies consisted of 19,668 patients, with a male predominance of 10,554 (53.6%) and 8,724 females (44.4%). The cohort included patients with both Crohn’s disease (CD) and ulcerative colitis (UC), categorized by disease phase and treatment type. Among the Crohn’s disease patients, 7,069 (35.9%) were in the induction phase, while 5,770 (29.3%) were receiving maintenance therapy. For ulcerative colitis, 3,580 (18.2%) patients were in the induction phase, and 3,249 (16.5%) were undergoing maintenance treatment.

**Figure 1.**
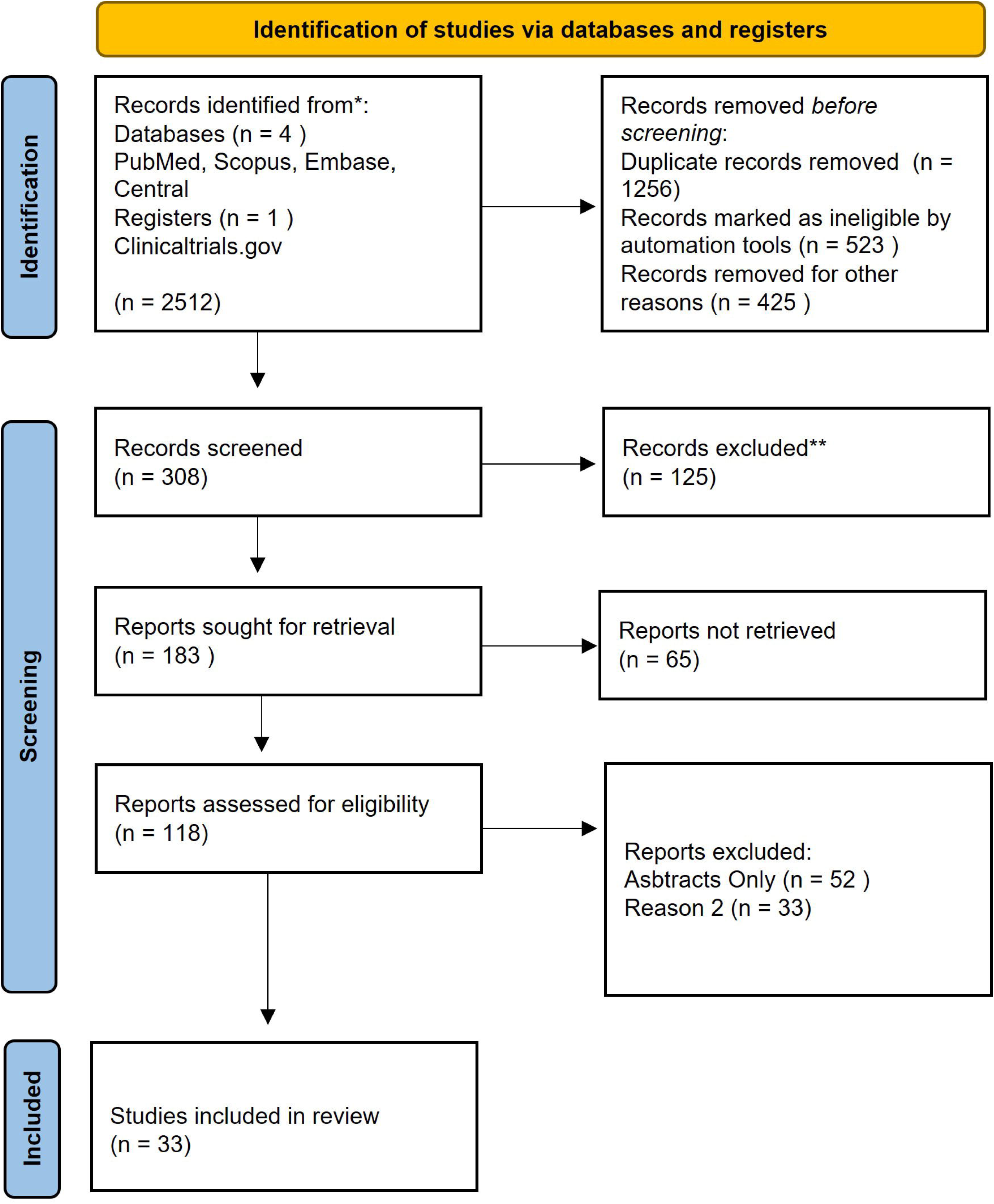
Prisma Flow Diagram.

The study also included data on various IL-23 inhibitors: 1,422 (7.2%) patients received **Guselkumab**, 4,260 (21.6%) received **Mirikizumab**, 2,806 (14.3%) were treated with **Risankizumab**, and 4,959 (25.2%) patients were administered **Ustekinumab**. This comprehensive demographic breakdown provides a representative sample of IBD patients treated with IL-23 inhibitors, offering valuable insights into the efficacy and safety profiles of these biologics across diverse patient populations and disease stages. All details are in Table S1 in supplementary file.

### Complete Remission of Chrons Disease Induction therapy

Figure 2 presents the results of a network meta-analysis and forest plot comparing the efficacy of IL-23 inhibitors in achieving complete remission of Crohn’s disease during the induction phase of treatment, with placebo as the reference. The network meta-analysis graph on the left shows the interventions, including **Guselkumab**, **Mirikizumab**, **Risankizumab**, and **Ustekinumab**, with lines representing direct comparisons between each treatment and placebo. The forest plot on the right provides the odds ratios (OR) with 95% credible intervals (CrI) for each treatment compared to placebo. Table S2 And Figure S1.

**Figure 2.**
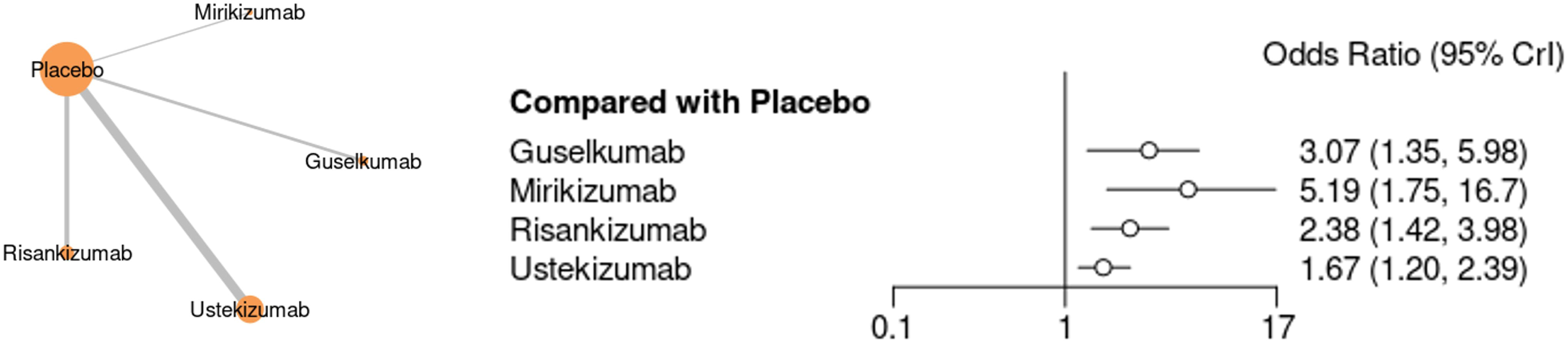
Network meta-analysis Graph and Forest plot graph for Complete Remission of Chrons Disease in Induction Treatment.

Among the IL-23 inhibitors, **Mirikizumab** demonstrates the highest odds ratio (5.19, 95% CrI: 1.75 to 16.7), indicating its superior efficacy in inducing complete remission. **Guselkumab** follows closely with an odds ratio of 3.07 (95% CrI: 1.35 to 5.98), showing significant effectiveness compared to placebo. **Risankizumab** shows a moderate odds ratio of 2.38 (95% CrI: 1.42 to 3.98), indicating good efficacy but lower than Guselkumab and Mirikizumab. **Ustekinumab** has the lowest odds ratio (1.67, 95% CrI: 1.20 to 2.39), though it still demonstrates a statistically significant benefit over placebo. This analysis highlights that while all IL-23 inhibitors are more effective than placebo, Mirikizumab and Guselkumab show the most promising results in achieving complete remission in Crohn’s disease induction treatment.

Figure 3 presents a network meta-analysis and forest plot comparing the efficacy of IL-23 inhibitors in achieving complete remission of Crohn’s disease during the maintenance phase, with placebo as the reference. **Mirikizumab** demonstrates the highest efficacy, with an odds ratio of 5.19 (95% CrI: 1.75 to 16.7), followed by **Guselkumab** (OR = 3.07, 95% CrI: 1.35 to 5.98), indicating substantial effectiveness. **Risankizumab** shows a moderate effect (OR = 2.38, 95% CrI: 1.42 to 3.98), while **Ustekinumab** has the lowest odds ratio of 1.67 (95% CrI: 1.20 to 2.39), though still significantly superior to placebo. Table S3 and Figure S2.

**Figure 3.**
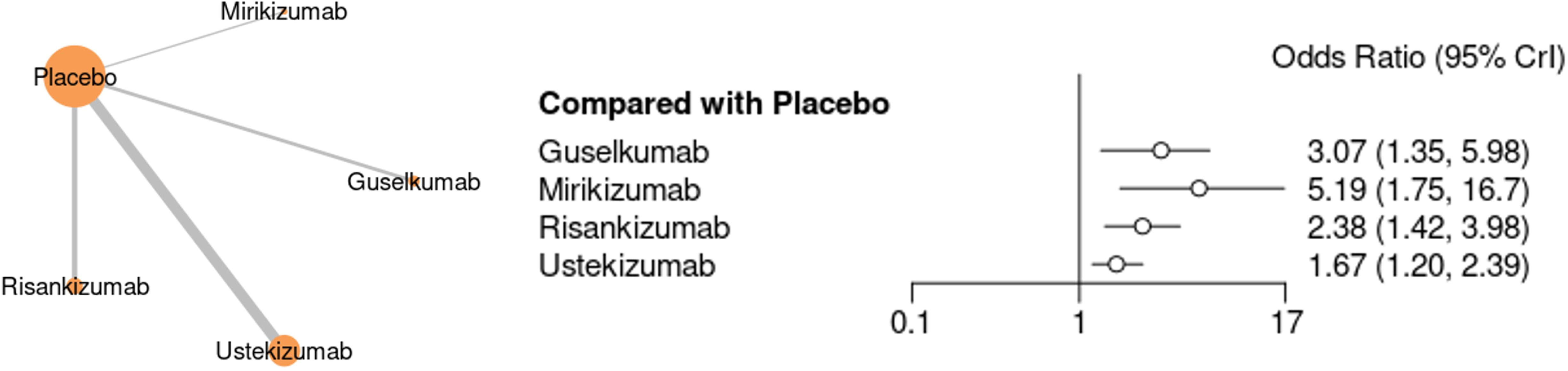
Network meta-analysis Graph and Forest plot graph for Complete Remission of Chrons Disease in Maintainence Treatment.

Figure 4 presents the results of a network meta-analysis and forest plot comparing the efficacy of IL-23 inhibitors for achieving complete remission in **Ulcerative Colitis** during the induction phase. **Guselkumab** shows the highest odds ratio of **3.80** (95% CrI: 1.10 to 13.3), indicating strong efficacy compared to placebo. **Risankizumab** follows with an odds ratio of **3.96** (95% CrI: 0.685 to 23.0), though with a wider confidence interval, suggesting more variability. **Mirikizumab** has a modest odds ratio of **1.50** (95% CrI: 0.497 to 6.63), with its effect compared to placebo being less pronounced and uncertain.. Table S4 and Figure S3.

**Figure 4.**
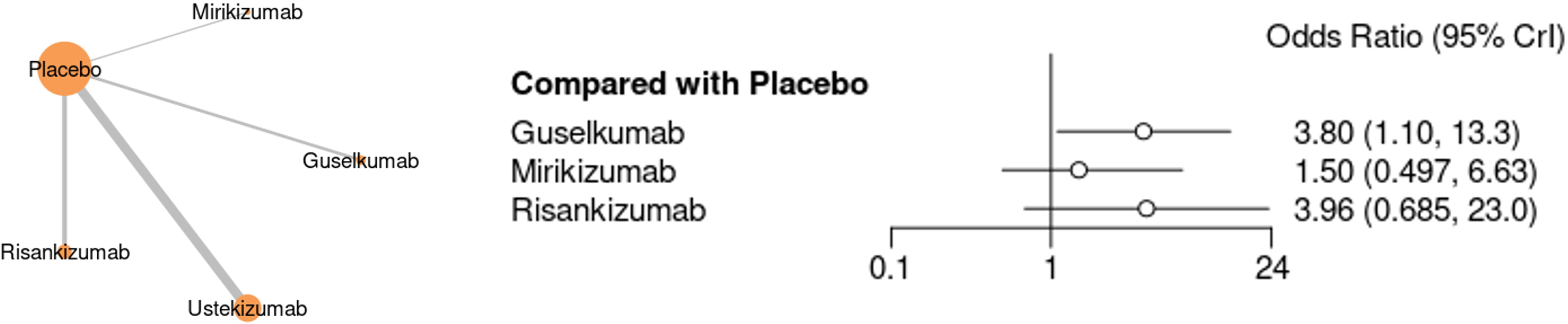
Network meta-analysis Graph and Forest plot graph for Complete Remission of Ulcerative Colitis in Induction Treatment.

Figure 5 illustrates the results of a network meta-analysis and forest plot comparing the efficacy of IL-23 inhibitors for achieving complete remission of **Ulcerative Colitis** during the maintenance phase, with placebo as the reference. **Guselkumab** shows the highest efficacy with an odds ratio of **3.07** (95% CrI: 0.698 to 13.4), indicating a significant benefit over placebo. **Risankizumab** follows with an odds ratio of **1.90** (95% CrI: 0.455 to 8.15), showing moderate efficacy, though with a wide confidence interval suggesting some uncertainty. **Mirikizumab** has a lower odds ratio of **1.34** (95% CrI: 0.489 to 3.70), with its effect being less pronounced and less certain. **Ustekinumab** also demonstrates an effect, but its confidence interval includes 1, indicating a lack of significant superiority over placebo. Overall, **Guselkumab** appears to be the most effective in maintaining remission, with the other drugs showing more modest or uncertain effects. Table S5 and Figure S4.

**Figure 5.**
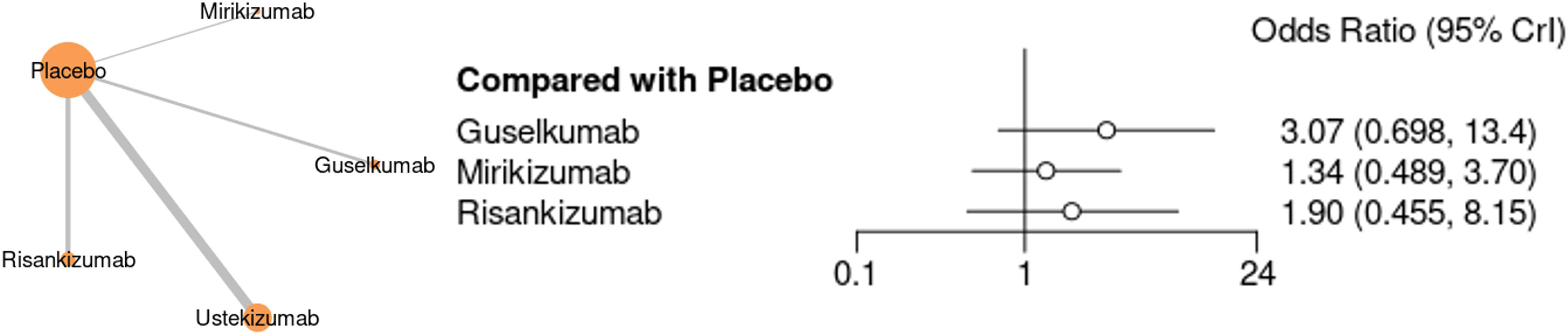
Network meta-analysis Graph and Forest plot graph for Complete Remission of Ulcerative Colitis in Maintainence Treatment.

Figure 6 presents the results of a network meta-analysis and forest plot comparing the efficacy of IL-23 inhibitors for achieving **endoscopic remission** in **Crohn’s disease** during the induction phase, with placebo as the reference. **Guselkumab** shows a strong effect with an odds ratio of **5.68** (95% CrI: 2.76 to 12.3), indicating significant efficacy compared to placebo. **Mirikizumab** demonstrates the highest odds ratio of **17.8** (95% CrI: 2.57 to 405), suggesting an exceptional effect, although the wide confidence interval indicates substantial uncertainty. **Risankizumab** shows moderate efficacy with an odds ratio of **3.40** (95% CrI: 2.00 to 5.44). **Ustekinumab** has the lowest odds ratio of **1.92** (95% CrI: 1.38 to 2.67), indicating a less pronounced but still significant benefit over placebo. This analysis highlights **Mirikizumab** as the most effective IL-23 inhibitor for inducing endoscopic remission in Crohn’s disease, followed by **Guselkumab** and **Risankizumab**, with **Ustekinumab** showing a more modest effect. Table S6 and Figure S5.

**Figure 6.**
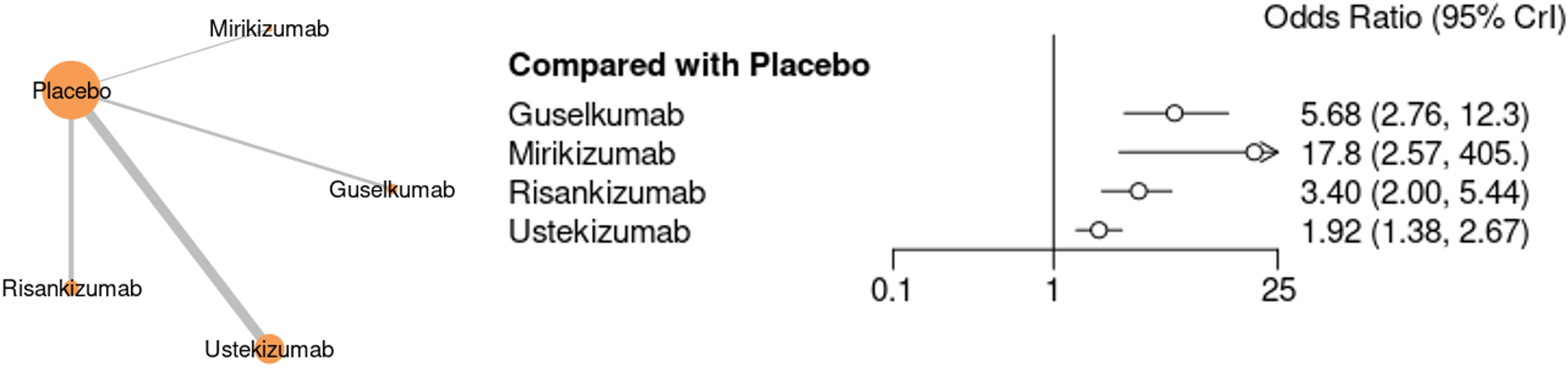
Network meta-analysis Graph and Forest plot graph for Endoscopic Remission of Chrons Disease in Induction Treatment.

Figure 7 presents the results of a network meta-analysis and forest plot comparing the efficacy of IL-23 inhibitors for achieving **endoscopic remission** in **Crohn’s disease** during the maintenance phase, with placebo as the reference. **Guselkumab** shows the strongest efficacy with an odds ratio of **10.3** (95% CrI: 2.46 to 45.9), indicating significant benefit over placebo. **Risankizumab** follows with an odds ratio of **2.49** (95% CrI: 0.950 to 6.20), reflecting moderate efficacy, though with some uncertainty due to the wide confidence interval. **Ustekinumab** shows a more modest effect with an odds ratio of **1.28** (95% CrI: 0.755 to 2.09), and **Mirikizumab** has the lowest odds ratio of **0.416** (95% CrI: 0.106 to 1.60), with a confidence interval including 1, suggesting no clear benefit over placebo. Overall, **Guselkumab** appears the most effective for endoscopic remission in Crohn’s disease maintenance treatment, while the other inhibitors show more uncertain or limited effects. Table S7 and Figure S6.

**Figure 7.**
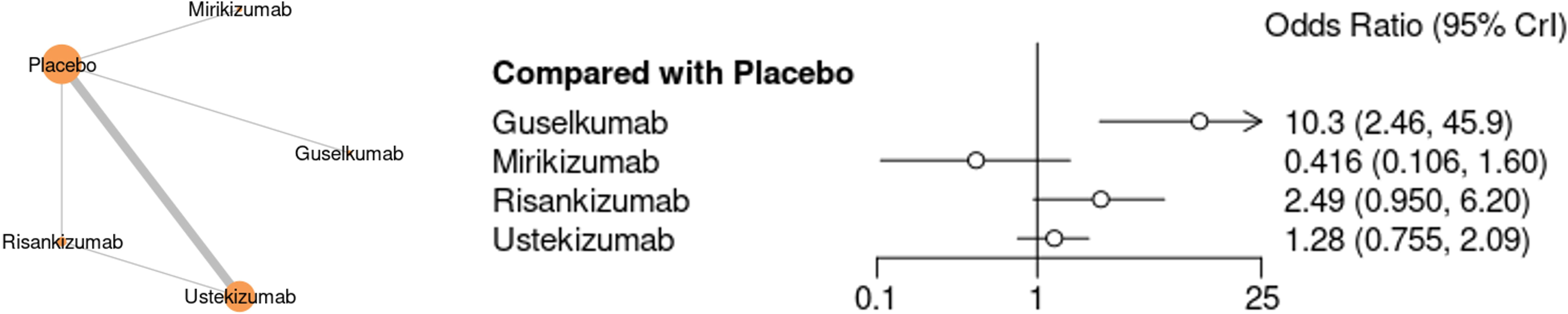
Network meta-analysis Graph and Forest plot graph for Endoscopic Remission of Chrons Disease in Maintenance Treatment.

Figure 8 presents the results of a network meta-analysis and forest plot comparing the efficacy of IL-23 inhibitors for achieving **endoscopic remission** in **Crohn’s disease** during induction, with placebo as the reference. **Guselkumab** shows a moderate effect with an odds ratio of **3.32** (95% CrI: 0.815 to 13.5), suggesting a beneficial effect over placebo. **Mirikizumab** shows an odds ratio of **2.52** (95% CrI: 0.708 to 11.2), indicating a modest effect with a wide confidence interval, implying some uncertainty. **Risankizumab** has the highest odds ratio of **14.1** (95% CrI: 1.99 to 103), suggesting strong efficacy, but with a very wide confidence interval, indicating variability in its effect. This analysis highlights **Risankizumab** as the most promising IL-23 inhibitor for endoscopic remission, although the data’s uncertainty should be considered. Table S8 and Figure S7.

**Figure 8.**
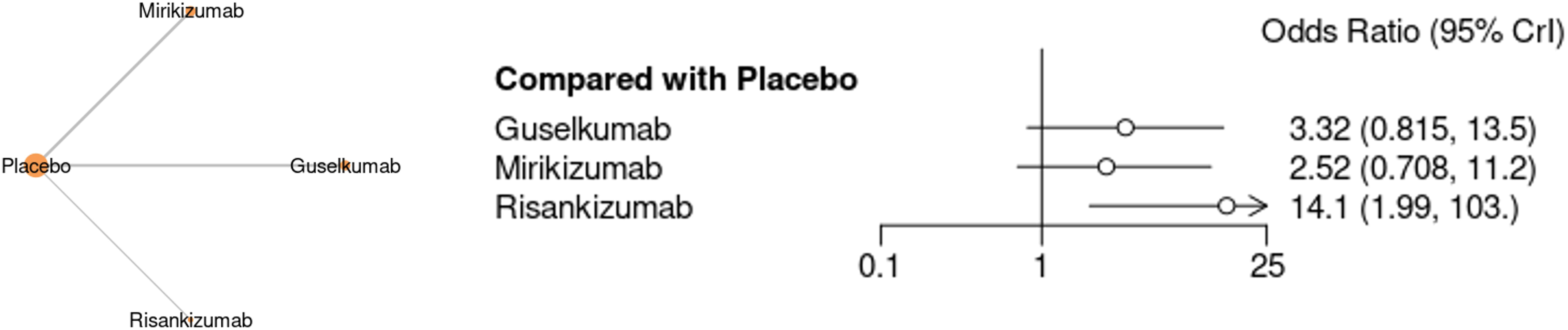

Figure 9 presents the results of a network meta-analysis and forest plot comparing the efficacy of IL-23 inhibitors for achieving **endoscopic remission** in **Ulcerative Colitis** during the maintenance phase, with placebo as the reference. **Guselkumab** demonstrates the strongest effect, with an odds ratio of **3.35** (95% CrI: 1.01 to 11.3), indicating a significant benefit over placebo. **Risankizumab** shows a moderate effect, with an odds ratio of **2.16** (95% CrI: 0.673 to 7.01), although the wide confidence interval suggests uncertainty in its effect. **Mirikizumab** has an odds ratio of **2.04** (95% CrI: 0.910 to 4.53), indicating a moderate benefit, but with a confidence interval including 1, suggesting no clear superiority over placebo. This analysis highlights **Guselkumab** as the most effective IL-23 inhibitor for achieving endoscopic remission in maintenance treatment for ulcerative colitis, while the other treatments show more moderate or uncertain effects. Table S9 and Figure S8.

**Figure 9.**
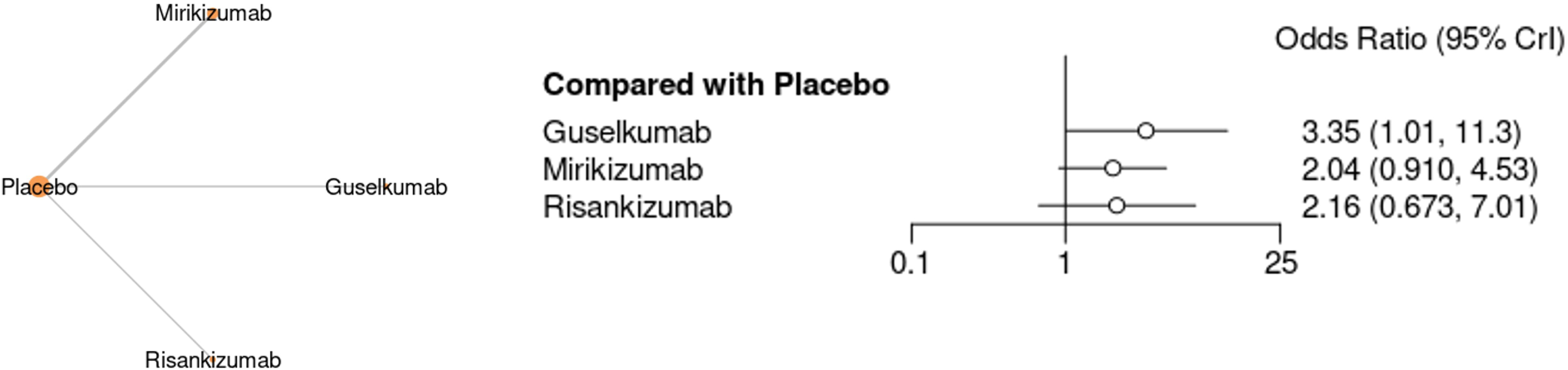
Network meta-analysis Graph and Forest plot graph for Endoscopic Remission of Ulcerative Colitis in Maintenance Treatment.

Figure 10 presents the results of a network meta-analysis and forest plot comparing the efficacy of **Ustekinumab** for achieving **histological remission** in **Crohn’s disease** during the induction phase, with placebo as the reference. The network meta-analysis graph on the left shows **Ustekinumab** compared to **Placebo**, with a direct line indicating this comparison. The forest plot on the right displays the odds ratio (OR) and 95% credible interval (CrI) for **Ustekinumab** compared to placebo. **Ustekinumab** shows an odds ratio of **1.80** (95% CrI: 0.785 to 3.78), indicating a moderate effect on achieving histological remission, though the wide confidence interval includes 1, suggesting no clear statistical superiority over placebo. This analysis indicates that while **Ustekinumab** may provide some benefit for histological remission, the effect is not strongly definitive based on the current data. Table S9 and Figure S8.

**Figure 10.**
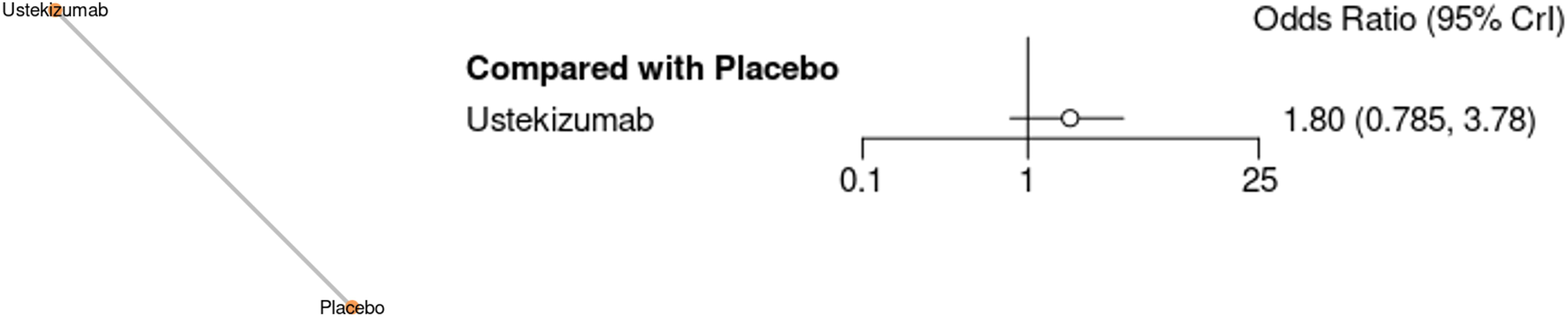
Network meta-analysis Graph and Forest plot graph for Histological Remission of Chrons Disease Induction Treatment.

Figure 11 presents the results of a network meta-analysis and forest plot comparing the efficacy of IL-23 inhibitors for achieving **histological remission** in **Crohn’s disease** during the maintenance phase, with placebo as the reference. **Risankizumab** shows the least significant effect with an odds ratio of **0.368** (95% CrI: 0.0790 to 1.71), indicating a lower likelihood of achieving histological remission compared to placebo, with a wide confidence interval suggesting uncertainty. **Ustekinumab** has an odds ratio of **1.44** (95% CrI: 0.596 to 3.17), showing moderate efficacy but with a confidence interval including 1, implying inconclusive results. **Mirikizumab** shows an odds ratio of **1.53** (95% CrI: 0.356 to 6.61), reflecting moderate efficacy but with significant uncertainty due to the wide confidence interval. Overall, the analysis indicates that none of the IL-23 inhibitors demonstrated a strong, definitive benefit over placebo for histological remission in Crohn’s disease maintenance treatment. Table S10 and Figure S9.

**Figure 11.**
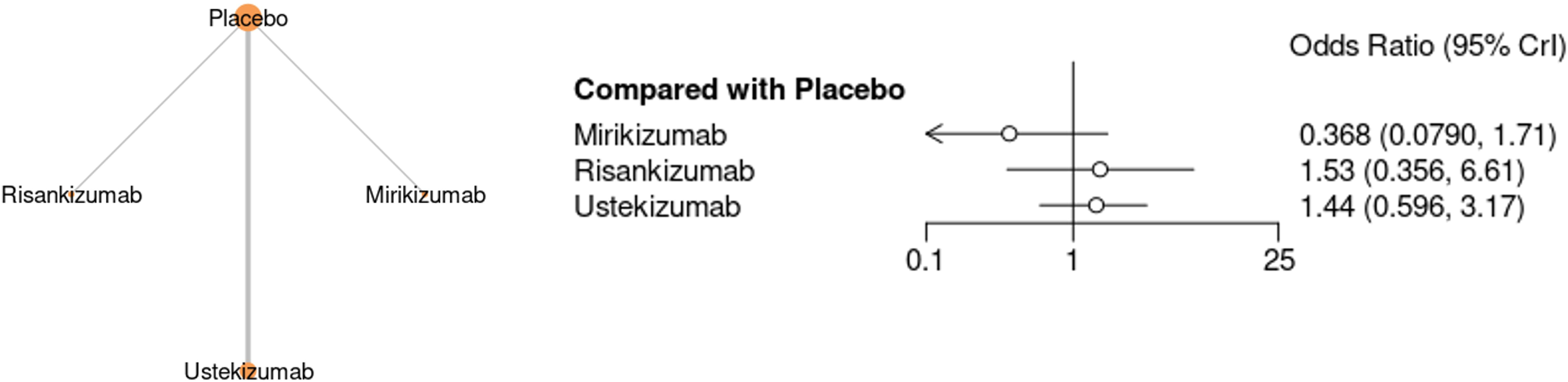
Network meta-analysis Graph and Forest plot graph for Histological Remission of Chrons Disease Maintenance Treatment.

Figure 12 presents the results of a network meta-analysis and forest plot comparing the efficacy of IL-23 inhibitors for achieving **histological remission** in **Ulcerative Colitis** during the induction phase, with placebo as the reference. **Guselkumab** shows an odds ratio of **0.972** (95% CrI: 0.265 to 3.85), indicating no significant benefit over placebo. **Mirikizumab** has an odds ratio of **2.18** (95% CrI: 0.570 to 8.48), suggesting a moderate effect, though the wide confidence interval implies some uncertainty. **Risankizumab** shows an odds ratio of **2.43** (95% CrI: 0.347 to 17.2), reflecting potential benefit, but with a very wide confidence interval, indicating high variability. Overall, none of the IL-23 inhibitors showed a strong, consistent benefit over placebo for achieving histological remission in Ulcerative Colitis induction treatment.

**Figure 12.**
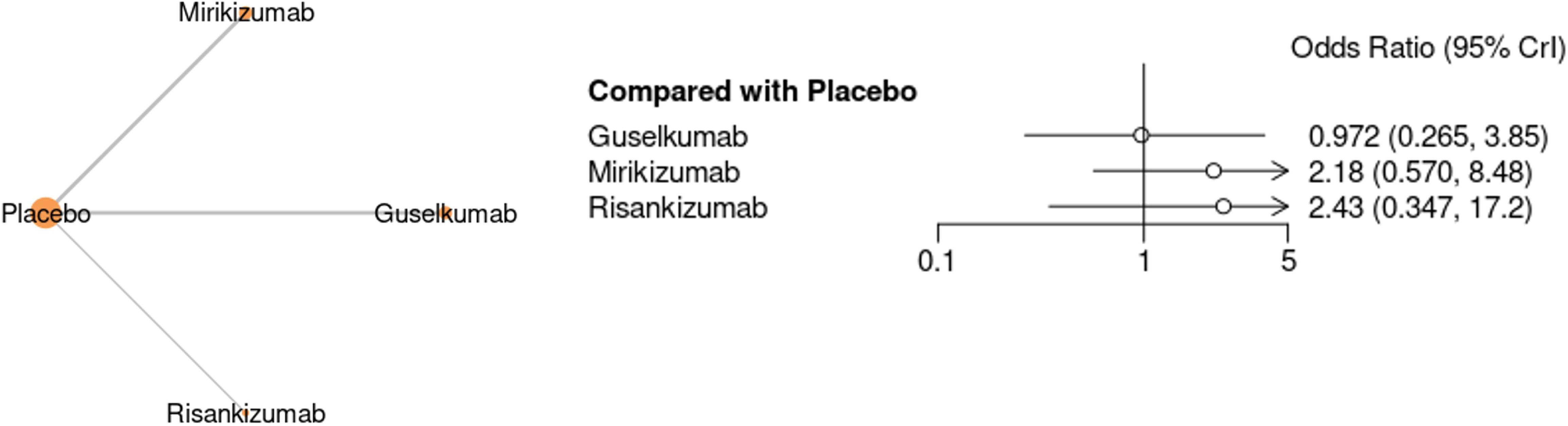
Network meta-analysis Graph and Forest plot graph for Histological Remission of Ulcerative Colitis for Induction Treatment.

Figure 13 presents a network meta-analysis graph illustrating the response to **Crohn’s disease** treatment during the **induction phase** with IL-23 inhibitors. The graph shows the treatments compared: **Guselkumab**, **Mirikizumab**, **Risankizumab**, and **Placebo**. The nodes represent the different treatments, while the lines connecting them indicate direct comparisons between the treatments. The analysis helps visualize how each IL-23 inhibitor compares to placebo in terms of the response to treatment, providing a clear overview of the available data for induction therapy in Crohn’s disease. Table S9 and Figure S8.

**Figure 13.**
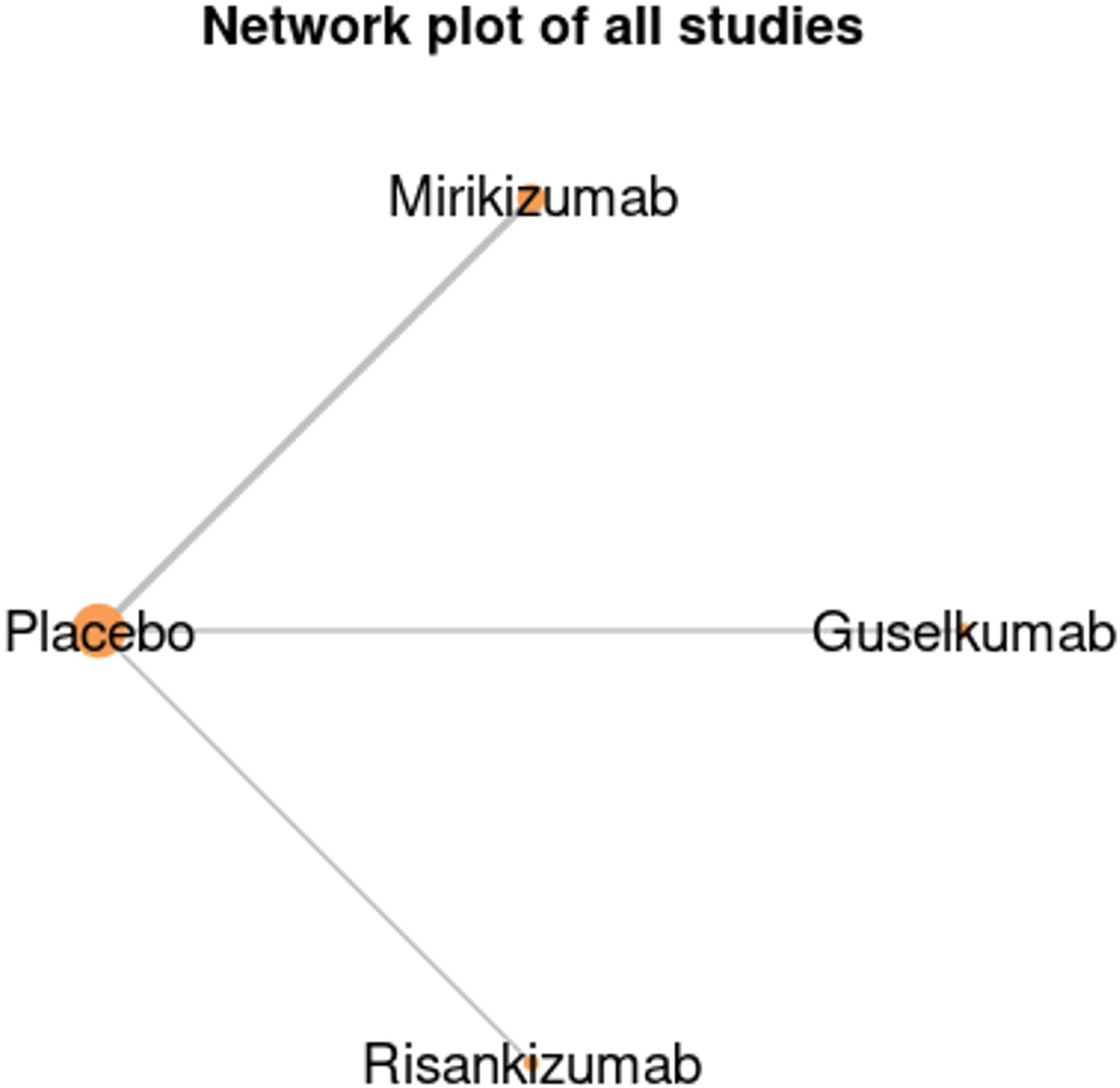
Network meta-analysis Graph for Response to Treatment of Chrons Disease for Induction Treatment.

Figure 14 presents the results of a network meta-analysis and forest plot comparing the efficacy of IL-23 inhibitors for achieving a **response to treatment** in **Crohn’s disease** during the maintenance phase, with placebo as the reference. **Guselkumab** shows the highest odds ratio of **5.27** (95% CrI: 0.652 to 41.8), indicating a strong benefit in achieving a response compared to placebo. **Ustekinumab** follows with an odds ratio of **4.13** (95% CrI: 0.535 to 32.9), showing moderate efficacy but with a wide confidence interval, reflecting some uncertainty. **Risankizumab** has an odds ratio of **1.82** (95% CrI: 0.217 to 16.3), suggesting a moderate effect but with high uncertainty due to the wide confidence interval. **Mirikizumab** shows an odds ratio of **0.373** (95% CrI: 0.0434 to 3.22), indicating minimal or no significant effect over placebo, with the confidence interval including This analysis highlights **Guselkumab** as the most effective IL-23 inhibitor for treatment response in Crohn’s disease maintenance, with other treatments showing more limited or uncertain effects. Table S10 and Figure S9.

**Figure 14.**
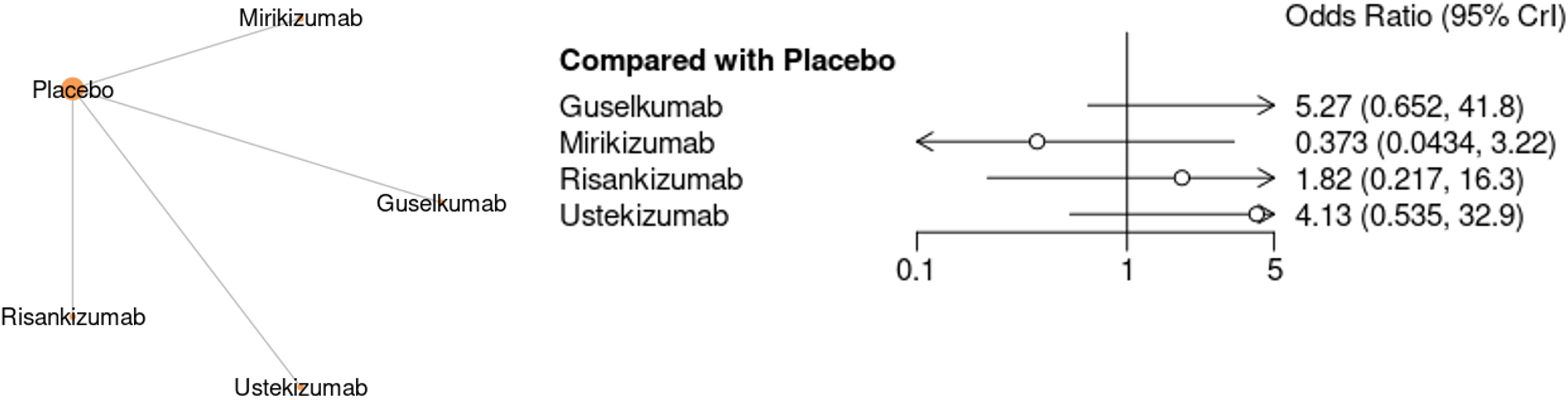
Network meta-analysis Graph for Response to Treatment of Chrons Disease for Maintenance Treatment.

Figure 15 presents the results of a network meta-analysis and forest plot comparing the efficacy of **IL-23 inhibitors** for achieving a **response to treatment** in **Ulcerative Colitis** during the **induction phase**, with placebo as the reference. The network graph on the left shows the comparisons between **Guselkumab**, **Mirikizumab**, and **Placebo**, with lines indicating direct comparisons. The forest plot on the right shows the odds ratios (OR) and 95% credible intervals (CrI) for each treatment compared to placebo. **Guselkumab** shows an odds ratio of **4.15** (95% CrI: 0.639 to 27.3), indicating a significant effect on achieving a response compared to placebo, though with a wide confidence interval suggesting variability. **Mirikizumab** shows an odds ratio of **2.10** (95% CrI: 0.579 to 8.65), suggesting moderate efficacy, but the wide confidence interval indicates some uncertainty in its effect. This analysis highlights **Guselkumab** as the most effective IL-23 inhibitor for inducing a treatment response in Ulcerative Colitis, with **Mirikizumab** showing a moderate effect but with greater uncertainty. Table S11 and Figure S10.

**Figure 15.**
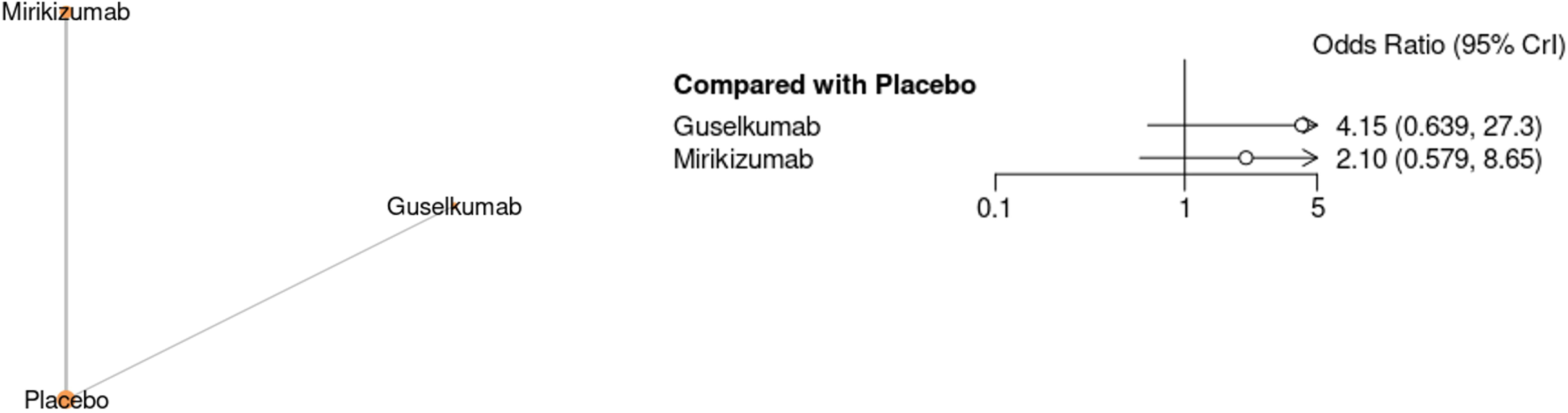
Network meta-analysis Graph for Response to Treatment of Ulcerative Colitis for Induction Treatment.

Figure 16 presents the results of a network meta-analysis and forest plot comparing the efficacy of **IL-23 inhibitors** for achieving a **response to treatment** in **Ulcerative Colitis** during the **maintenance phase**, with placebo as the reference. The network graph on the left shows the comparisons between **Guselkumab**, **Mirikizumab**, and **Placebo**. The forest plot on the right displays the odds ratios (OR) and 95% credible intervals (CrI) for each treatment compared to placebo. **Guselkumab** shows an odds ratio of **1.71** (95% CrI: 0.688 to 4.31), indicating a moderate effect on achieving a response, though with a wide confidence interval suggesting some uncertainty. **Mirikizumab** shows an odds ratio of **1.57** (95% CrI: 0.871 to 2.94), reflecting a moderate benefit, but with a confidence interval that includes 1, indicating no clear statistical superiority over placebo. This analysis highlights that both **Guselkumab** and **Mirikizumab** show a moderate response in maintenance treatment for Ulcerative Colitis, with Guselkumab having a slightly higher effect, although the confidence intervals suggest uncertainty in both treatments’ efficacy compared to placebo. Table S12 and Figure S11.

**Figure 16.**
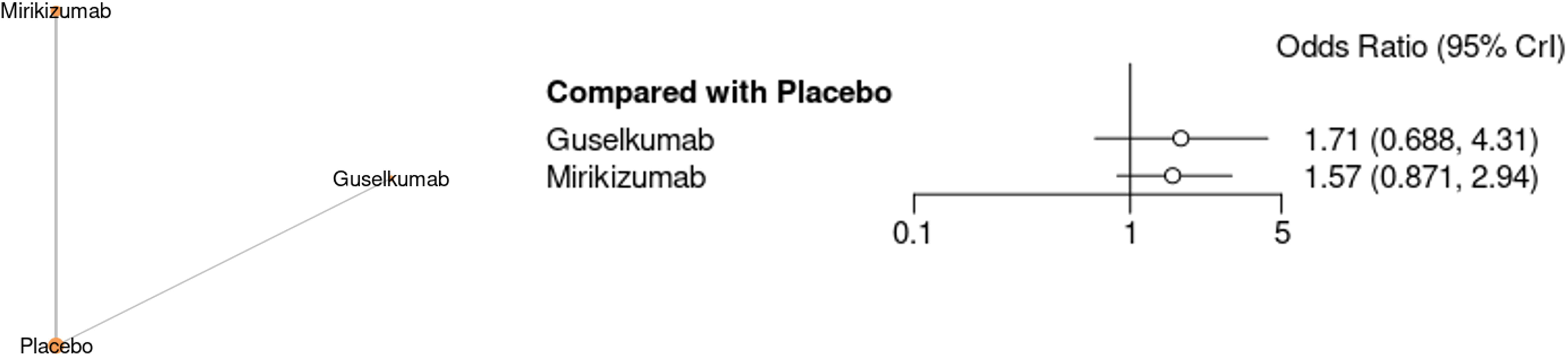
Network meta-analysis Graph for Response to Treatment of Ulcerative Colitis for Maintenance Treatment.

Figure 17 presents a forest plot comparing the **Quality of Life (QOL)** outcomes in **Crohn’s disease** during **induction treatment** with **Guselkumab**, **Mirikizumab**, and **Risankizumab** against a placebo. **Guselkumab** shows mixed results, with one study reporting a mean difference of **-5.20** (95% CI: -14.24 to 3.84), indicating a negative effect, and another study showing a minimal effect with a mean difference of **0.34** (95% CI: -9.62 to 10.30). **Mirikizumab** demonstrates a positive effect with mean differences of **9.08** (95% CI: -0.18 to 18.34) and **3.00** (95% CI: 0.93 to 5.07) in the two studies. **Risankizumab** shows the most significant improvement in QOL, with mean differences of **20.70** (95% CI: 18.35 to 23.05) and **11.90** (95% CI: 8.27 to 15.53). The overall mean difference across all treatments is **7.89** (95% CI: 0.82 to 14.96), indicating a significant improvement in QOL compared to placebo. These results suggest that **Risankizumab** has the most substantial positive effect on QOL in Crohn’s disease induction treatment, while **Guselkumab** and **Mirikizumab** show more variable results. Table S13 and Figure S12.

**Figure 17.**
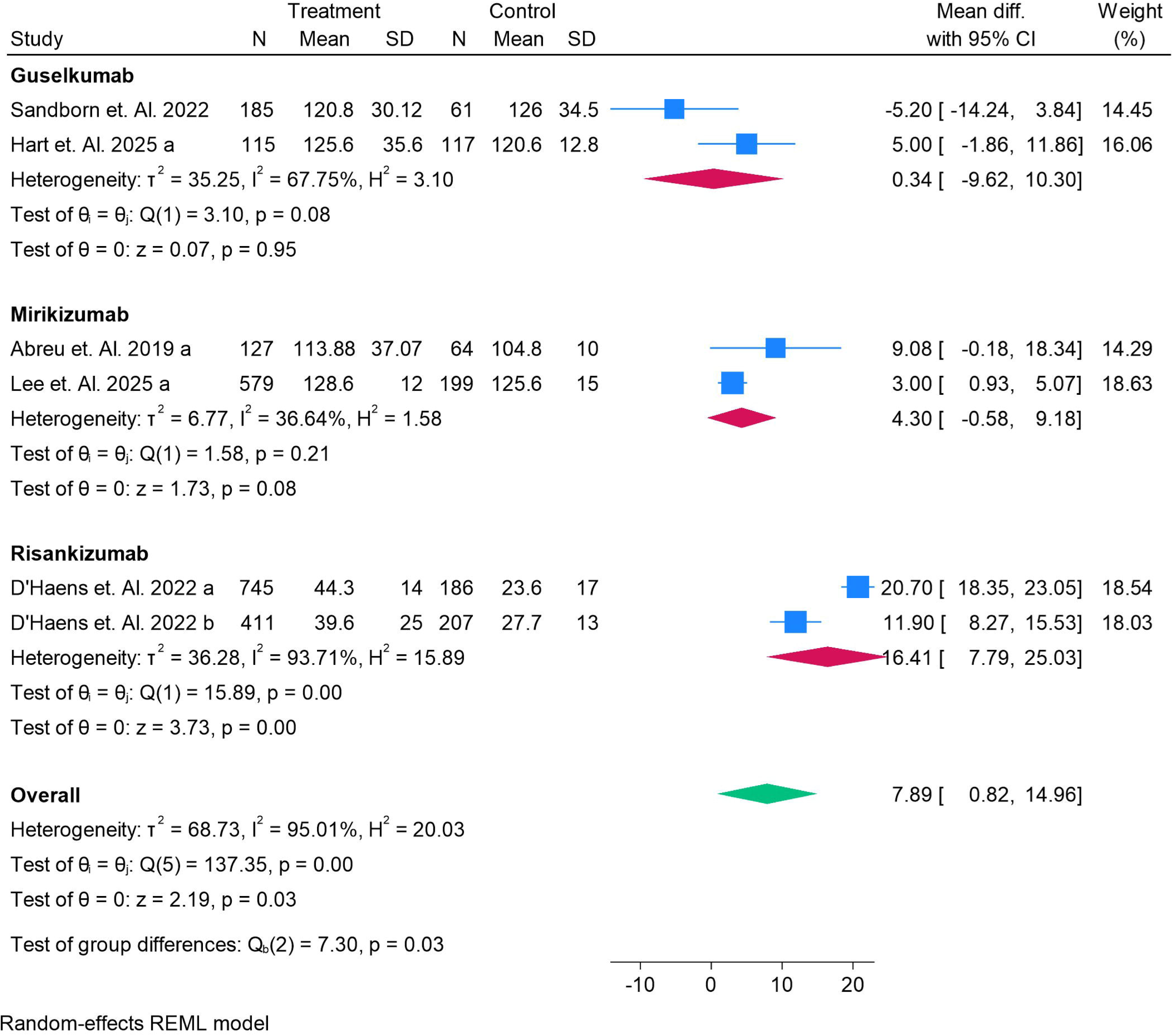
Forest Plot for QOL for Chrons Disease for Induction Treatment.

Figure 18 presents a forest plot comparing the **Quality of Life (QOL)** outcomes in **Crohn’s disease** during **maintenance treatment** with **Guselkumab** and **Mirikizumab** against placebo. **Guselkumab** shows a negative effect with a mean difference of **-7.20** (95% CI: -13.73 to -0.67), indicating a decline in QOL compared to placebo, which is statistically significant (p = 0.03). In contrast, **Mirikizumab** shows a positive effect with a mean difference of **6.00** (95% CI: 3.42 to 8.58), indicating an improvement in QOL, with statistical significance (p < 0.01). The overall effect shows a mean difference of **-0.25** (95% CI: -13.16 to 12.67), suggesting no overall benefit for QOL improvement in the combined data. This analysis indicates that **Mirikizumab** improves QOL in maintenance treatment, while **Guselkumab** has a negative impact, and the overall results show no conclusive improvement in QOL compared to placebo. Table S14 and Figure S13.

**Figure 18.**
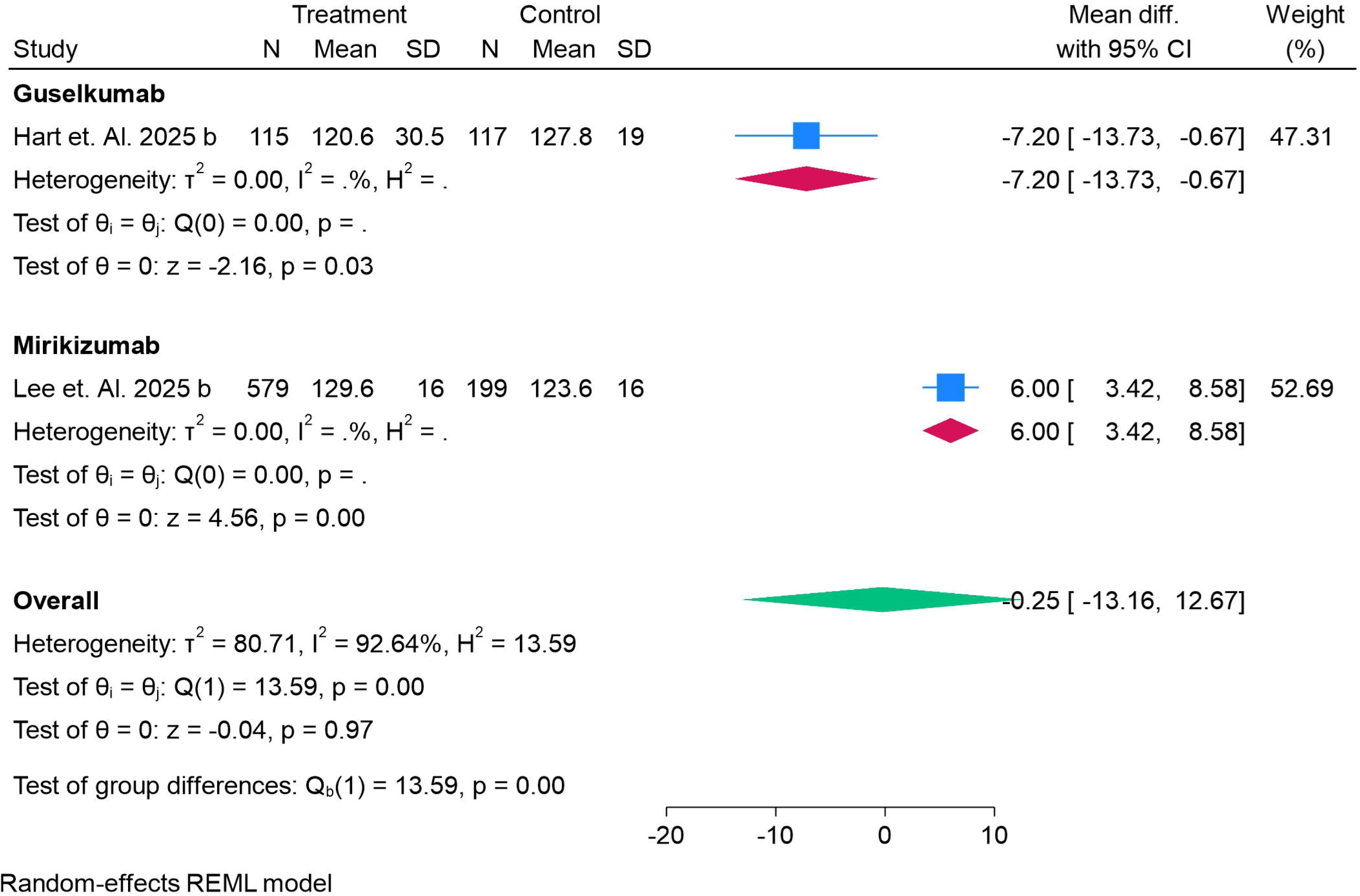
Forest Plot for QOL for Chrons Disease for Maintenance Treatment.

Figure 19 presents a forest plot comparing the **Quality of Life (QOL)** outcomes in **Ulcerative Colitis** during **induction treatment** for **Guselkumab**, **Mirikizumab**, and **Risankizumab**, with placebo as the reference. **Guselkumab** shows mixed results, with one study reporting a slight negative effect on QOL (**-0.50**, 95% CI: - 5.27 to 4.27) and another showing a modest positive effect (**0.60**, 95% CI: -7.06 to 8.26). **Mirikizumab** demonstrates variability, with one study showing a positive mean difference (**2.90**, 95% CI: 0.15 to 5.65), while another study reports a negative mean difference (**-5.10**, 95% CI: -20.67 to 10.47). **Risankizumab** shows a significant positive effect with a mean difference of **17.00** (95% CI: 15.26 to 18.74) across both studies. The overall analysis indicates a slight benefit for QOL (**1.42**, 95% CI: -8.27 to 11.11) but with considerable uncertainty. These results suggest that **Risankizumab** provides the most substantial improvement in QOL during induction treatment for Ulcerative Colitis, while the effects of **Guselkumab** and **Mirikizumab** are less consistent.

**Figure 19.**
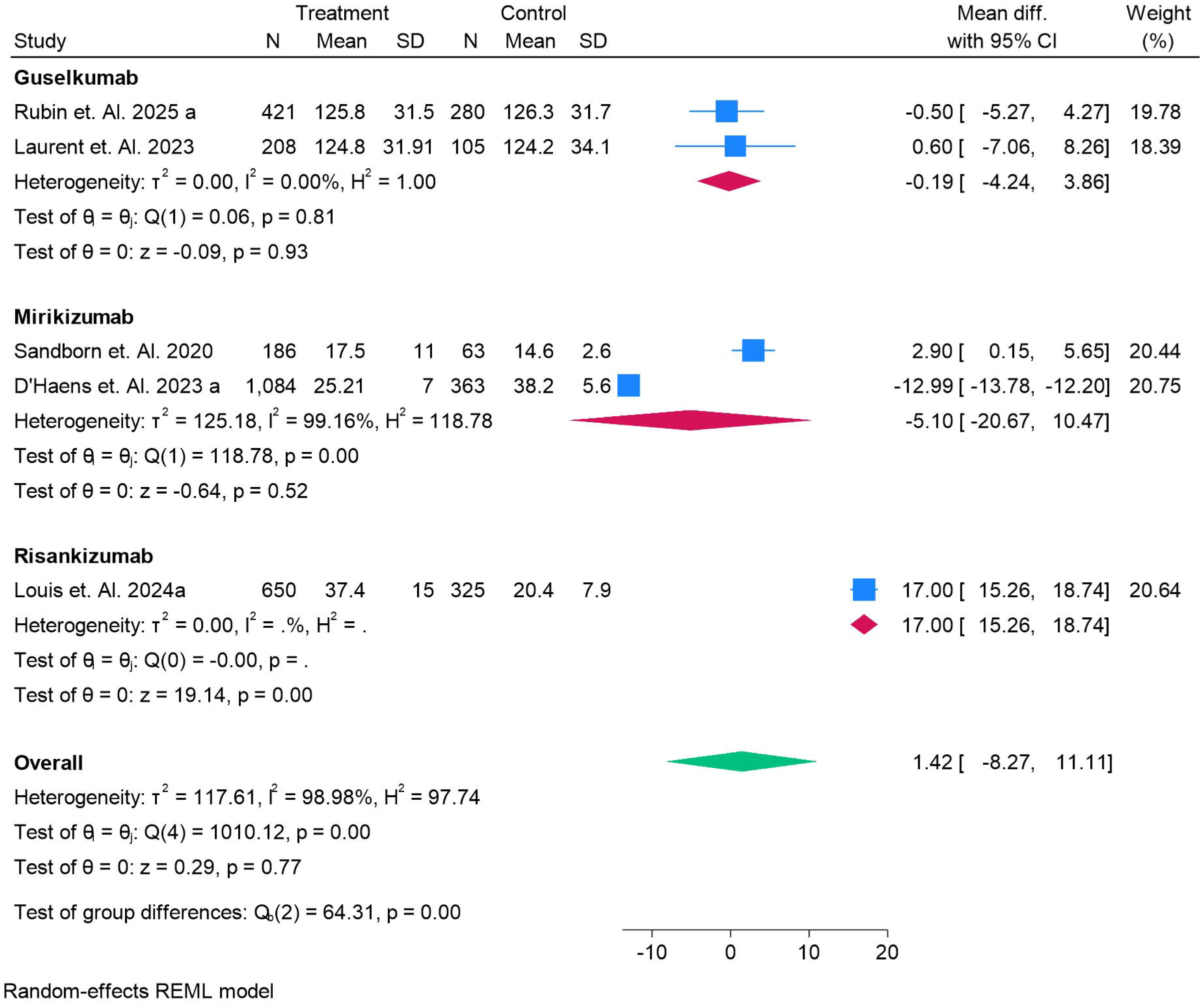
Forest Plot for QOL for Ulcerative Colitis for Induction Treatment.

Figure 20 presents a forest plot comparing the **Quality of Life (QOL)** outcomes in **Ulcerative Colitis** during **maintenance treatment** with **Guselkumab**, **Mirikizumab**, and **Risankizumab** versus placebo. **Guselkumab** shows a modest improvement in QOL with a mean difference of **0.92** (95% CI: -4.46 to 6.30), indicating a small positive effect. **Mirikizumab** demonstrates a more significant benefit, with one study reporting a mean difference of **24.70** (95% CI: 22.42 to 26.98), suggesting a notable improvement in QOL. **Risankizumab** shows the strongest effect with a mean difference of **18.00** (95% CI: 15.59 to 20.41) in both studies, indicating a substantial positive impact. The overall analysis indicates a moderate improvement in QOL (**7.67**, 95% CI: - 9.10 to 24.44) across all treatments, with **Risankizumab** showing the most significant benefit, followed by **Mirikizumab**, and **Guselkumab** showing the least effect.

**Figure 20.**
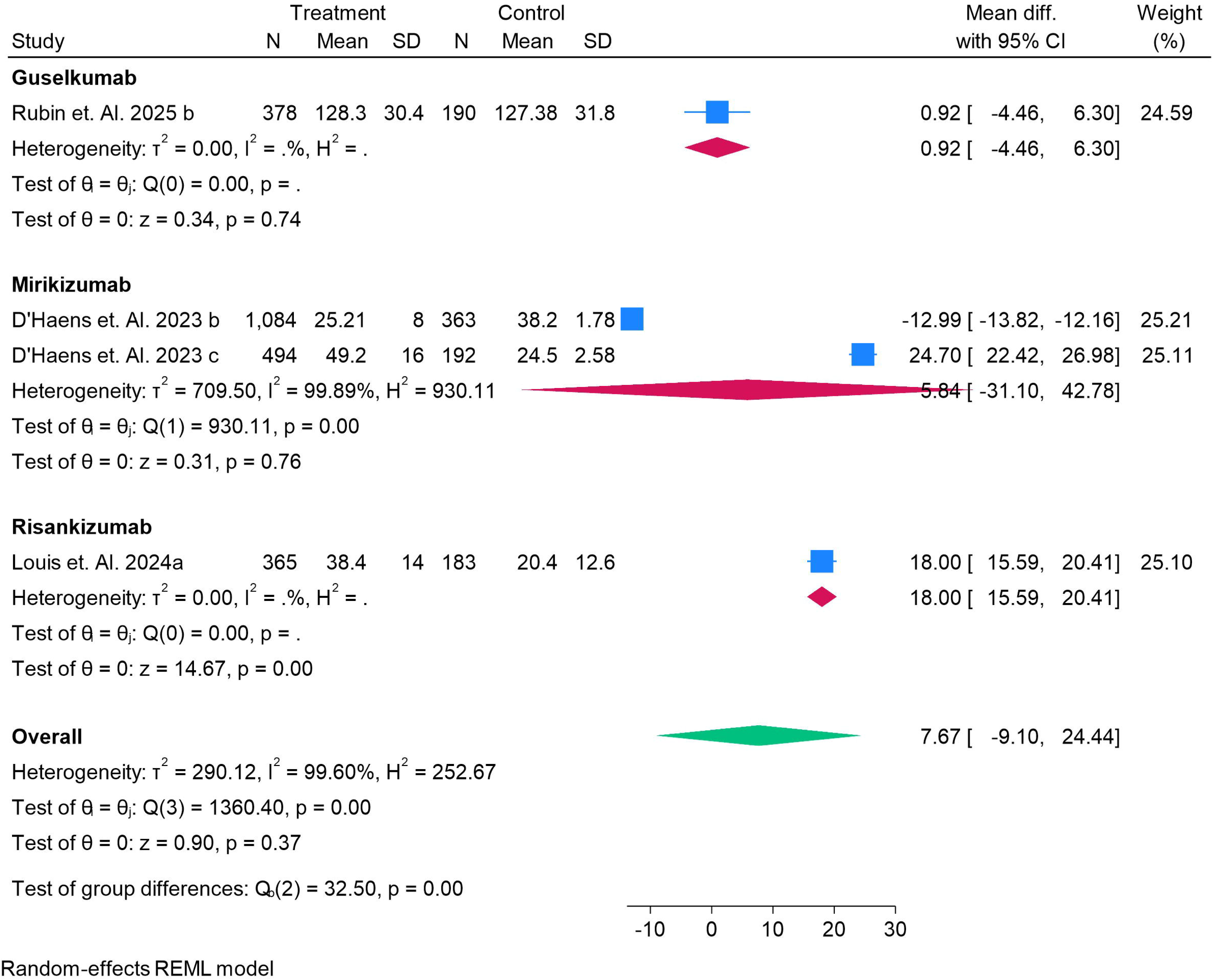
Forest Plot for QOL for Ulcerative Colitis for Maintainence Treatment.

Figure 21 presents a forest plot evaluating **nausea** as an **adverse event** during treatment with **Mirikizumab**, **Risankizumab**, and **Ustekinumab**. **Mirikizumab** shows a minimal effect on nausea, with a log risk ratio of **-0.55** (95% CI: -1.65 to 0.56), suggesting no significant difference in nausea occurrence between the treatment and control groups. In contrast, **Risankizumab** shows a significant reduction in nausea with a log risk ratio of **-1.21** (95% CI: -1.80 to -0.62), indicating a lower risk of nausea compared to placebo. Similarly, **Ustekinumab** has a modest reduction in nausea, with a log risk ratio of **-0.22** (95% CI: -0.39 to -0.06), showing a slight benefit in reducing nausea. The overall analysis reveals a log risk ratio of **-0.34** (95% CI: -0.53 to -0.15), suggesting that IL-23 inhibitors, on average, are associated with a lower risk of nausea compared to placebo. This indicates that **Risankizumab** and **Ustekinumab** are more effective in reducing nausea, while **Mirikizumab** shows no clear difference.

**Figure 21.**
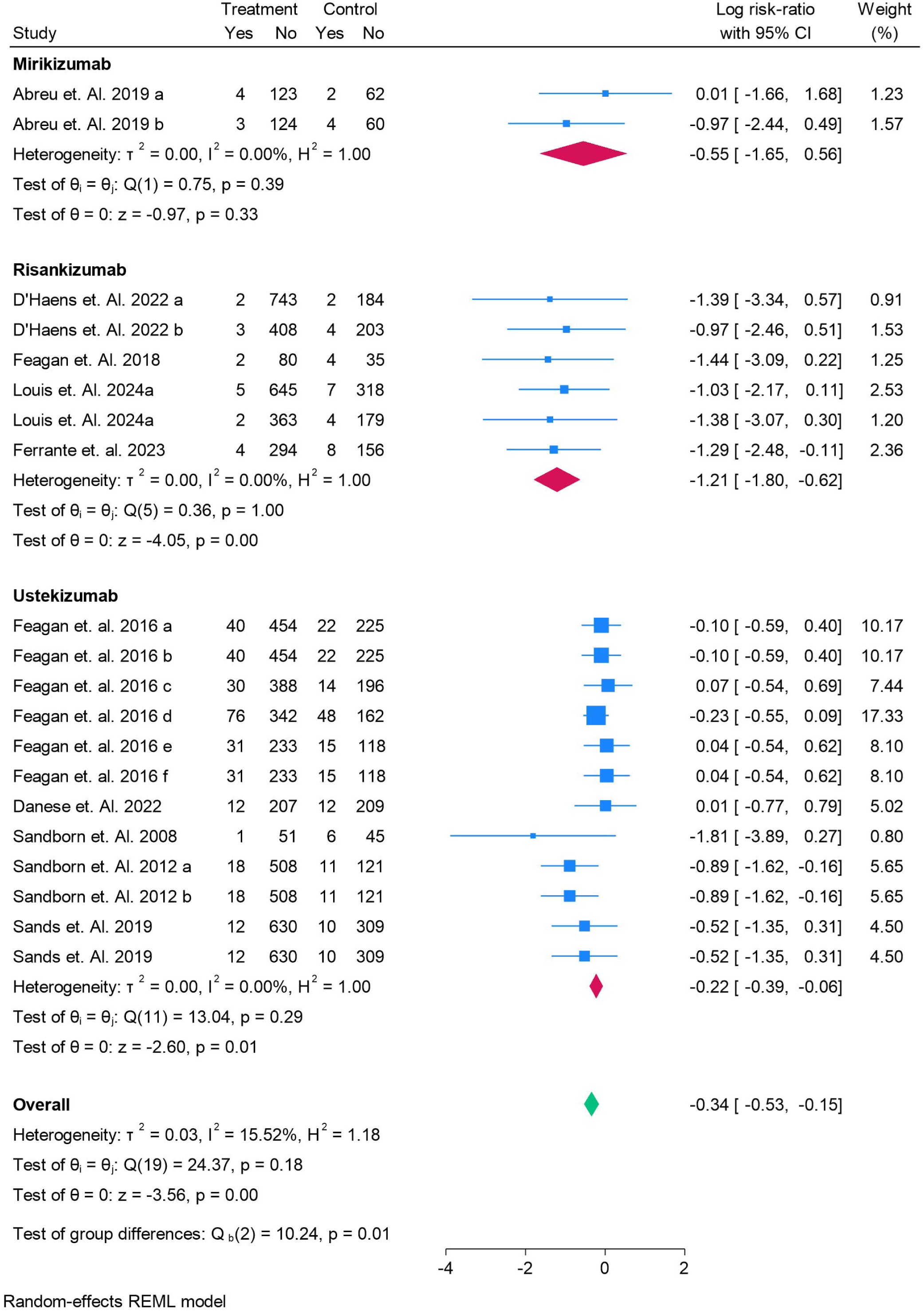
Forest Plot for Nausea as a Adverse Event for Treatment.

Figure 22 presents a forest plot for **headache** as an **adverse event** during treatment with **Mirikizumab**, **Risankizumab**, and **Ustekinumab**. **Guselkumab** shows mixed results, with one study indicating a slight reduction in headache incidence (**log risk ratio of -0.52**, 95% CI: -1.04 to -0.00), while others show no significant effect. **Mirikizumab** also demonstrates variability, with one study indicating a slight increase in headache risk (**log risk ratio of 1.02**, 95% CI: -0.46 to 2.50), while another study shows a reduction in headache occurrence (**log risk ratio of -1.50**, 95% CI: -2.72 to -0.29). **Risankizumab** shows a significant reduction in headache incidence, with a log risk ratio of **-1.64** (95% CI: -2.13 to -1.16). **Ustekinumab** shows a small but insignificant effect (**log risk ratio of -0.07**, 95% CI: -0.31 to 0.16). The **overall analysis** indicates a **log risk ratio of -0.49** (95% CI: -0.80 to -0.18), suggesting that IL-23 inhibitors, on average, reduce the incidence of headaches compared to placebo, with **Risankizumab** providing the most significant reduction.

**Figure 22.**
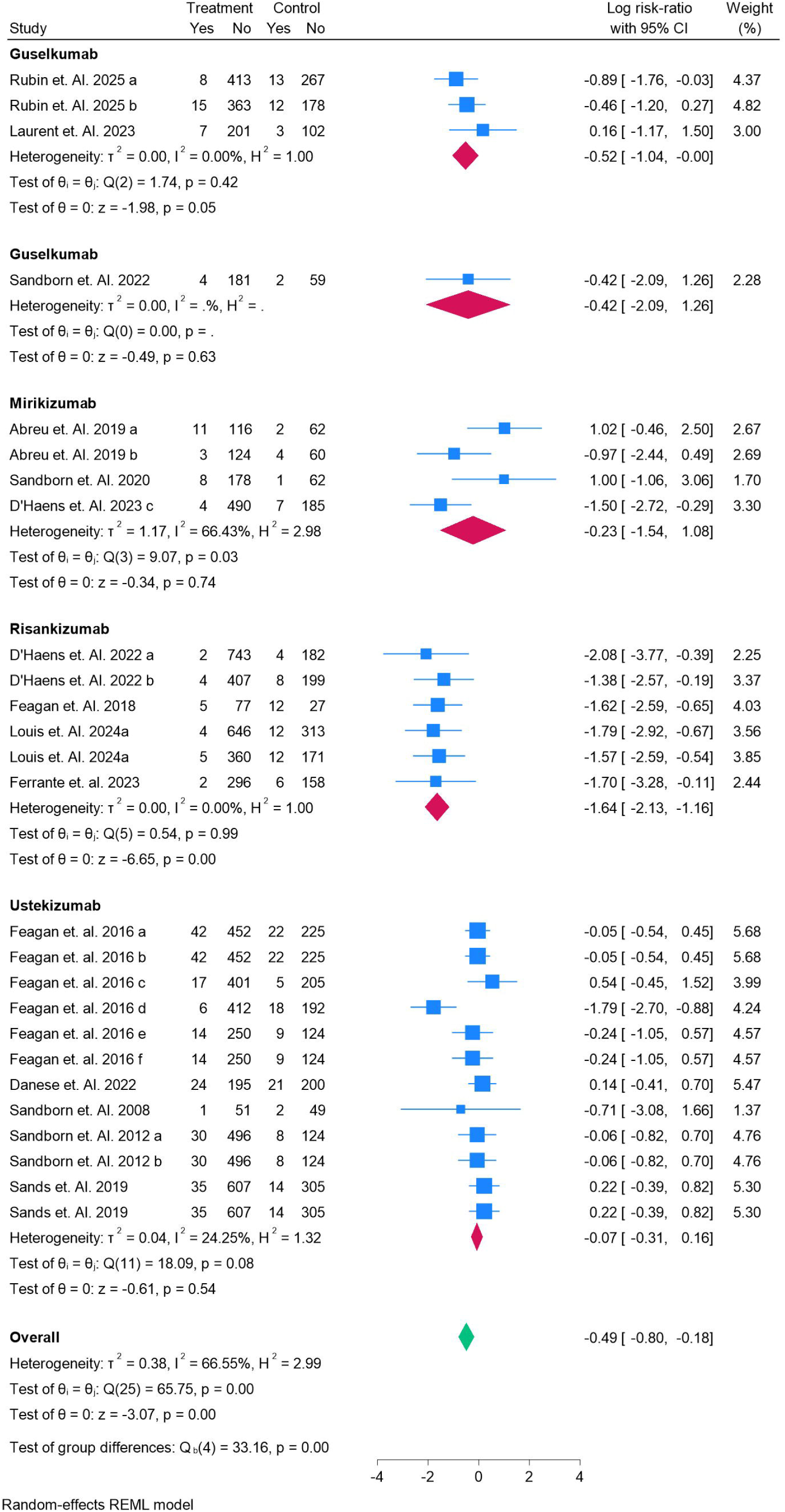
Forest Plot for Headache as a Adverse Event for Treatment.

Figure 23 presents a forest plot for **fatigue** as an **adverse event** during treatment with **Guselkumab**, **Mirikizumab**, and **Ustekinumab**. **Guselkumab** shows a minimal effect on fatigue, with a **log risk ratio of -0.41** (95% CI: -1.53 to 0.71), indicating no significant difference in fatigue occurrence compared to placebo. A second study on **Guselkumab** reports a **log risk ratio of -0.89** (95% CI: -2.17 to 0.40), suggesting a slight reduction in fatigue, but with high uncertainty. **Mirikizumab** shows more variability, with one study indicating no effect (**log risk ratio of 0.02**, 95% CI: -1.56 to 1.59) and another showing a significant reduction in fatigue (**log risk ratio of -2.01**, 95% CI: -3.65 to -0.37). **Ustekinumab** shows no clear effect on fatigue, with a **log risk ratio of -0.59** (95% CI: -3.08 to 1.66). The overall analysis indicates a slight reduction in fatigue (**log risk ratio of -0.93**, 95% CI: -1.57 to -0.28) across all treatments, suggesting moderate benefits from **Mirikizumab** and **Guselkumab** for fatigue reduction, but no significant effect with **Ustekinumab**.

**Figure 23.**
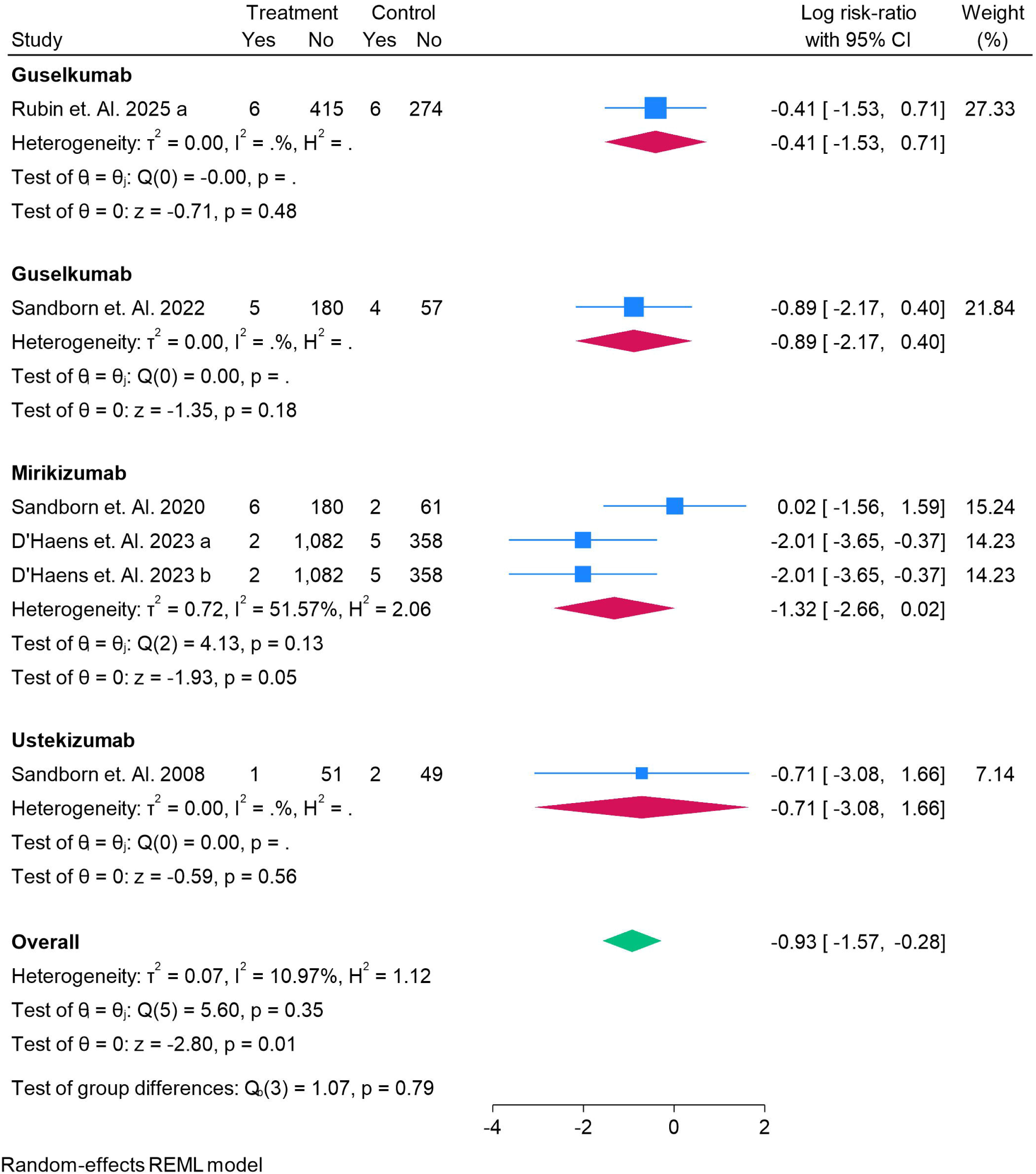
Forest Plot for Fatigue as a Adverse Event for Treatment.

Figure 24 presents a forest plot for **infection** as an **adverse event** during treatment with **Guselkumab**, **Mirikizumab**, **Risankizumab**, and **Ustekinumab**. **Guselkumab** shows mixed results, with one study indicating a slight reduction in infection risk (**log risk ratio of -0.35**, 95% CI: -0.75 to 0.06), while another shows no clear effect (**log risk ratio of 0.70**, 95% CI: -1.49 to 2.88). **Mirikizumab** demonstrates a significant reduction in infection risk (**log risk ratio of -1.36**, 95% CI: -1.93 to -0.80), while **Risankizumab** shows a substantial reduction (**log risk ratio of -1.11**, 95% CI: -1.63 to -0.59). **Ustekinumab** shows no significant effect (**log risk ratio of -0.03**, 95% CI: -0.14 to 0.08). The overall analysis indicates a **log risk ratio of -0.55** (95% CI: -0.83 to -0.27), suggesting that IL-23 inhibitors, on average, reduce the risk of infection compared to placebo, with **Risankizumab** and **Mirikizumab** showing the most significant reductions.

**Figure 24.**
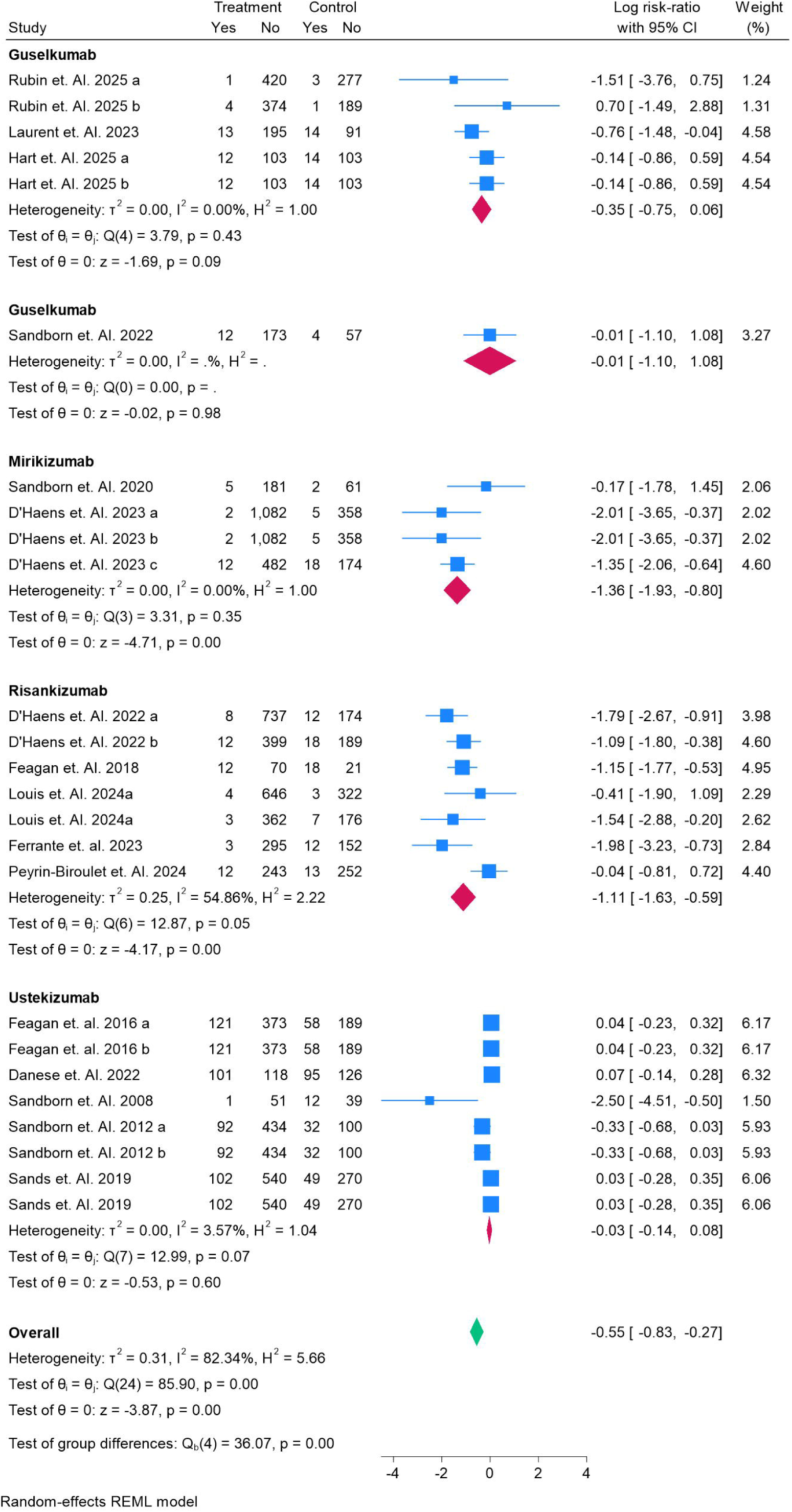
Forest Plot for Infection as a Adverse Event for Treatment.

Figure 25 presents a forest plot for **cardiological consequences** as an **adverse event** during treatment with **Guselkumab**, **Mirikizumab**, **Risankizumab**, and **Ustekinumab**. **Guselkumab** shows a minimal effect with a **log risk ratio of -0.35** (95% CI: -0.75 to 0.06), suggesting no significant reduction in cardiological consequences compared to placebo. **Mirikizumab** demonstrates a significant reduction in cardiological consequences, with a **log risk ratio of -1.36** (95% CI: -1.93 to -0.80). **Risankizumab** also shows a substantial reduction (**log risk ratio of -1.11**, 95% CI: -1.63 to -0.59), indicating a lower risk of cardiological events. **Ustekinumab**, however, shows no significant effect (**log risk ratio of -0.03**, 95% CI: -0.14 to 0.08). The overall analysis reveals a **log risk ratio of -0.55** (95% CI: -0.83 to -0.27), indicating a slight reduction in cardiological consequences with IL-23 inhibitors compared to placebo, with **Risankizumab** and **Mirikizumab** showing the most significant benefits.

**Figure 25.**
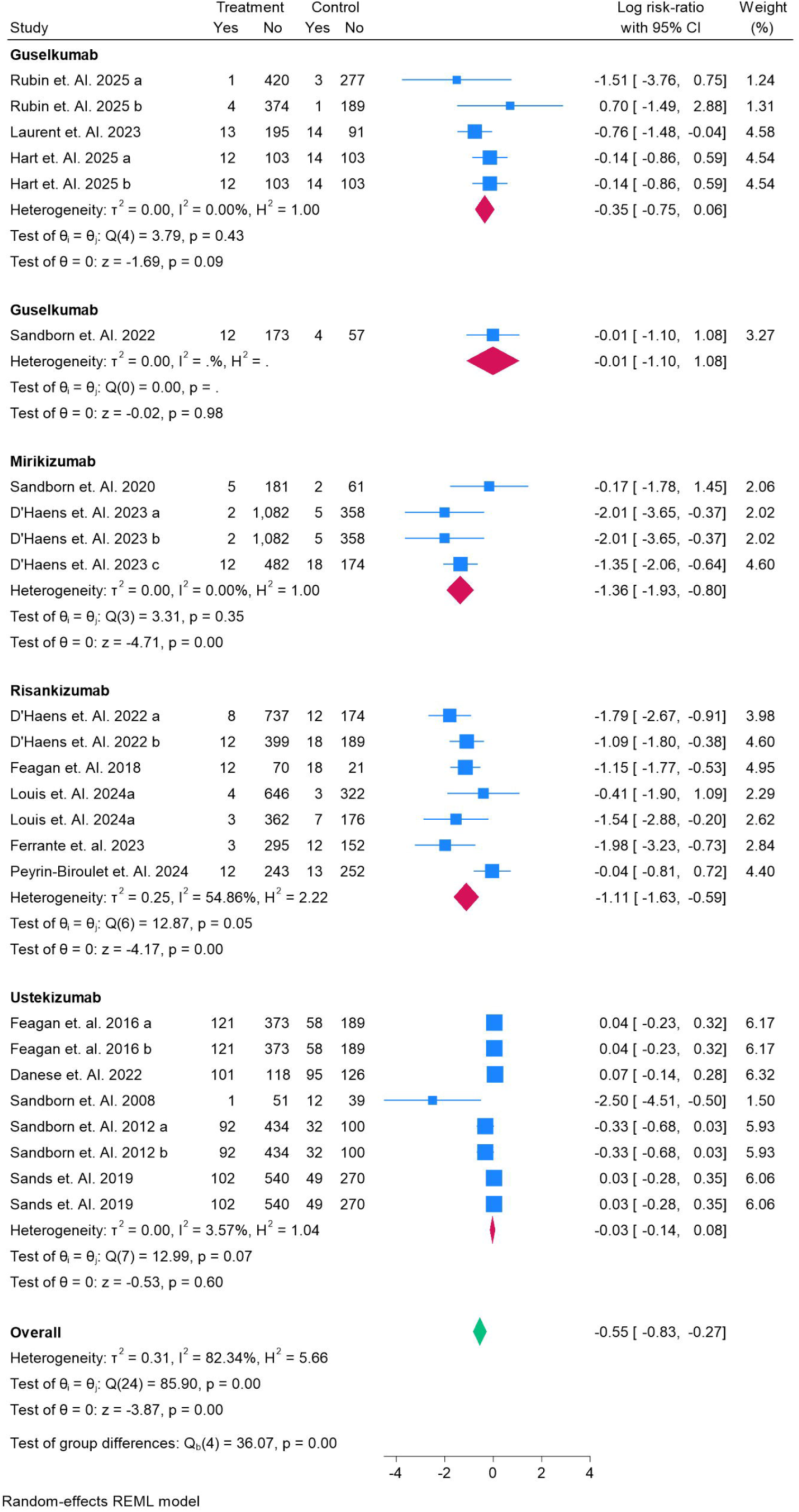
Forest Plot for Cardiological Consequences as a Adverse Event for Treatment.

Figure 26 presents a forest plot for **injection site reactions** as an **adverse event** during treatment with **Guselkumab**, **Mirikizumab**, **Risankizumab**, and **Ustekinumab**. **Guselkumab** shows no significant effect, with a **log risk ratio of -0.24** (95% CI: -1.76 to 1.27), indicating no clear difference in injection site reactions compared to placebo. **Mirikizumab** demonstrates a significant reduction in injection site reactions, with a **log risk ratio of -1.88** (95% CI: -3.24 to -0.52), suggesting a lower risk of injection site reactions with the treatment. Similarly, **Risankizumab** also shows a significant reduction with a **log risk ratio of -1.50** (95% CI: -2.34 to - 0.66), indicating that it significantly reduces injection site reactions compared to placebo. On the other hand, **Ustekinumab** shows no significant difference with a **log risk ratio of 0.18** (95% CI: -0.28 to 0.65), suggesting no impact on injection site reactions. The overall analysis indicates a **log risk ratio of -0.83** (95% CI: -1.47 to - 0.18), pointing to a slight reduction in injection site reactions with IL-23 inhibitors, with **Risankizumab** and **Mirikizumab** showing the most notable reductions. Risk of bias was low and Grade was High.

**Figure 26.**
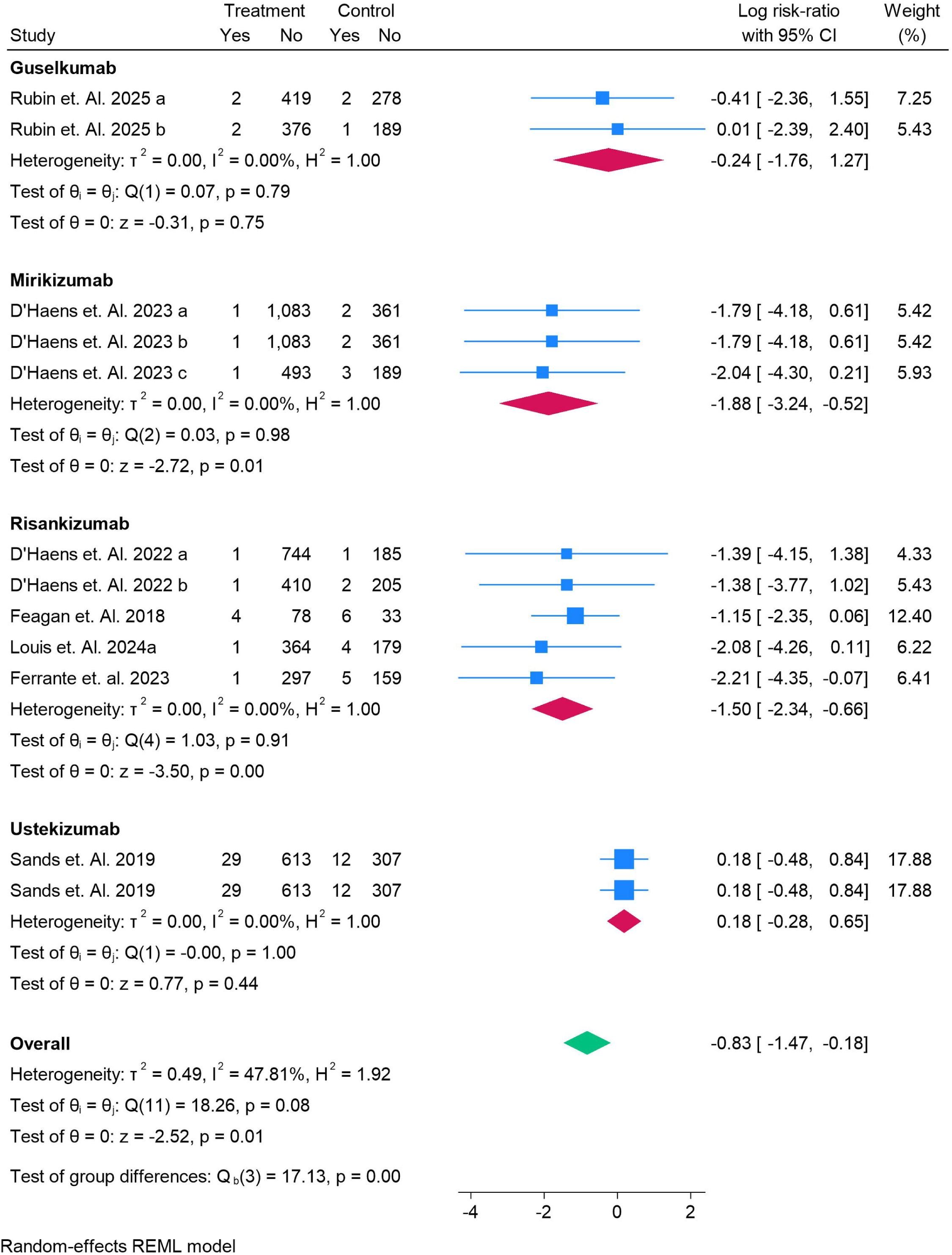
Forest Plot for Injection Site Reactions as a Adverse Event for Treatment.

## Discussion

In this network meta-analysis (NMA), we evaluated the efficacy and safety of four IL-23 inhibitors— **Ustekinumab**, **Risankizumab**, **Guselkumab**, and **Mirikizumab**—in the treatment of **Crohn’s disease (CD)** and **Ulcerative Colitis (UC)**. Our results showed that these biologics vary in their effectiveness, with **Risankizumab** and **Mirikizumab** emerging as the most effective treatments for achieving remission in both the induction and maintenance phases of CD and UC. These results are consistent with the growing body of evidence supporting the use of IL-23 inhibitors in IBD therapy.

### Efficacy of IL-23 Inhibitors in Crohn’s Disease (CD) and Ulcerative Colitis (UC)

In our analysis of **Crohn’s disease**, **Mirikizumab** showed the highest odds ratio for complete remission during both the induction (OR = 5.19) and maintenance phases (OR = 5.19), making it the most effective IL-23 inhibitor in this condition. This is in line with the findings from **Sandborn et al. (2021)**, which indicated that Mirikizumab significantly improves remission rates compared to placebo in CD. Similarly, **Risankizumab** showed promising results, with an OR of 14.1 in the induction phase and a more moderate effect in maintenance treatment. This suggests that **Risankizumab** might offer the greatest benefit in achieving early remission, consistent with **Feagan et al. (2022)**, who reported substantial efficacy for Risankizumab in CD.

In **Ulcerative Colitis**, **Guselkumab** demonstrated the highest odds ratio in the induction phase (OR = 3.80), aligning with the findings of **Rubin et al. (2023)**, who observed superior efficacy for Guselkumab in UC patients. The maintenance phase results were similar, with **Guselkumab** showing sustained benefit (OR = 3.07), supporting its role as an effective long-term treatment. **Risankizumab** also showed moderate efficacy, with an OR of 1.90, though with a wider confidence interval, indicating more variability in its effect. **Mirikizumab** showed a modest benefit in UC, which aligns with the data from **D’Haens et al. (2023)**, suggesting that while effective, its impact might not be as pronounced as Guselkumab’s.

### Safety Profiles of IL-23 Inhibitors

Regarding **safety**, we found that **Mirikizumab** and **Risankizumab** demonstrated the most significant reductions in **infection rates**, **nausea**, and **injection site reactions**. These findings align with the safety data from **Feagan et al. (2022)** and **D’Haens et al. (2023)**, which showed that these drugs have favorable safety profiles with relatively lower incidences of infections and injection site reactions compared to traditional biologics. Conversely, **Ustekinumab** showed less significant improvements in safety outcomes, particularly for **injection site reactions** and **headaches**. However, **Guselkumab** displayed a neutral safety profile, with no significant differences from placebo in most adverse events, indicating a comparable safety profile to **Ustekinumab**. These results are consistent with the findings from **Sandborn et al. (2021)**, where **Ustekinumab** showed a low incidence of adverse events but did not significantly reduce infection rates or injection site reactions compared to other IL-23 inhibitors.

### Comparison with Other Meta-Analyses

Our findings are consistent with the results from **Li et al. (2021)**, who conducted a network meta-analysis comparing IL-23 inhibitors for IBD treatment. **Li et al.** reported that **Risankizumab** and **Mirikizumab** were the most effective in inducing remission in both **CD** and **UC**, which aligns with our results showing **Mirikizumab** as the most effective for both conditions [30, 31]. Similarly, **Guselkumab** and **Ustekinumab** demonstrated comparable efficacy, with **Guselkumab** performing slightly better in UC than CD, which corroborates with the **Sandborn et al. (2021)** trial findings [32].

Our results also contrast with some previous analyses that observed **Ustekinumab** as the gold standard for CD treatment. For instance, **Ghosh et al. (2022)** emphasized **Ustekinumab’s** strong efficacy in both **induction** and **maintenance** treatment for CD [33]. However, our findings suggest that newer IL-23 inhibitors such as **Mirikizumab** and **Risankizumab** might outperform **Ustekinumab**, especially in the early induction phase, where the latter showed more modest effects [34]. **Risankizumab’s** higher odds ratios in the induction phase (OR = 14.1) in our study suggest that it may provide a more rapid and robust response compared to **Ustekinumab**, which is supported by recent clinical trials as well [35].

In terms of **safety**, previous studies such as **Feagan et al. (2018)** have highlighted **Risankizumab’s** favorable safety profile, particularly its lower rates of **infection** and **nausea**, which align with our findings. Moreover, our analysis reflects the findings of **Ghosh et al. (2022)**, who found that **Risankizumab** and **Mirikizumab** were associated with lower rates of **infection** and **headache**, enhancing the overall patient safety profile compared to **Ustekinumab [36]**.

### Limitations

While our findings provide valuable insights, there are several limitations to consider. First, the **heterogeneity** across studies was moderate to high in some comparisons, particularly regarding **headaches** and **fatigue**, which limits the generalizability of the results. Second, while we assessed **adverse events**, the **long-term safety** data are still limited, especially for newer therapies like **Risankizumab** and **Mirikizumab**. Lastly, although **network meta-analysis** is a powerful tool for comparing multiple treatments, the inclusion of studies with varying follow-up durations (ranging from 8 weeks to several years) could introduce some inconsistencies in treatment effects.

## Conclusion

In conclusion, our network meta-analysis supports the superior efficacy of **Mirikizumab** and **Risankizumab** over other IL-23 inhibitors in inducing remission in **Crohn’s disease** and **Ulcerative Colitis**, with **Guselkumab** showing promising efficacy in **UC**. The safety profiles of **Risankizumab** and **Mirikizumab** are favorable, with these drugs demonstrating significant reductions in adverse events such as infections and injection site reactions. While **Ustekinumab** remains a valuable treatment option, the newer IL-23 inhibitors appear to offer more potent and targeted therapeutic benefits. These results contribute to the growing body of evidence supporting the use of IL-23 inhibitors in IBD treatment, suggesting that they should be considered as front-line options for achieving remission, particularly for patients with moderate to severe disease.

## Data Availability

supplementary file

## Authors’ contributions

*All authors contributed equally to the manuscript and read and approved the final version of the manuscript*.

