## supplementary file for "Efficacy and Safety of IL-23 Inhibitors in the Treatment of Crohn’s Disease and Ulcerative Colitis: Systemic Review and Network Meta-Analysis"

Search String

("IL-23 inhibitors" OR "ustekinumab" OR "risankizumab" OR "guselkumab" OR "mirikizumab") AND ("Crohn's disease" OR "ulcerative colitis" OR "inflammatory bowel disease") AND ("efficacy" OR "remission" OR "mucosal healing" OR "clinical response") AND ("safety" OR "adverse events" OR "infection" OR "malignancy")

Table S1. Demographics and Summary Findings

| Ref No | **Author Year** | **Country** | **Disease** | **Therapy** | **Total Sample** | **Total Male** | **Total Female** | **Treatment Name** | **Treatment Sample** | **Control Sample** | **Control Number** | Grade | **Main Finding** |
| --- | --- | --- | --- | --- | --- | --- | --- | --- | --- | --- | --- | --- | --- |
| 12 | Rubin et. Al. 2025 a | Multicenter | Ulcerative Colities | Induction | 701 | 399 | 302 | Guselkumab | 421 | Placebo | 280 | High | **Guselkumab was effective and safe for both induction and maintenance of remission in patients with moderately to severely active ulcerative colitis.** |
| 12 | Rubin et. Al. 2025 b | Multicenter | Ulcerative Colities | Maintainence | 568 | 311 | 176 | Guselkumab | 378 | Placebo | 190 | High | **Guselkumab was effective and safe for both induction and maintenance of remission in patients with moderately to severely active ulcerative colitis.** |
| 13 | Sandborn et. Al. 2022 | Multicenter | Crohns Disease | Induction | 309 | 183 | 126 | Guselkumab | 185 | Placebo | 61 | High | **Guselkumab** demonstrated significantly greater clinical and endoscopic improvements in Crohn’s disease patients compared to placebo . |
| 14 | Laurent et. Al. 2023 | Multicenter | Ulcerative Colities | Induction | 208 | 119 | 66 | Guselkumab | 208 | Placebo | 105 | High | Guselkumab induction therapy was more effective than placebo in achieving clinical response, remission, and improvement in quality of life in patients with moderately to severely active ulcerative colitis. |
| 15 | Hart et. Al. 2025 a | **United Kingdom**, **Canada**, **Brazil**, **Italy**, **Japan**, **China**, **Germany**, and the **United States**. | Crohns Disease | Induction | 232 | 133 | 99 | Guselkumab | 115 | Placebo | 117 | High | Guselkumab significantly improved clinical remission and endoscopic response in participants with moderately to severely active Crohn’s disease when compared to placebo, with improved quality of life and fewer adverse events. |
| 15 | Hart et. Al. 2025 b | **United Kingdom**, **Canada**, **Brazil**, **Italy**, **Japan**, **China**, **Germany**, and the **United States**. | Crohns Disease | Maintainence | 232 | 135 | 97 | Guselkumab | 115 | Placebo | 117 | High | Guselkumab significantly improved clinical remission and endoscopic response in participants with moderately to severely active Crohn’s disease when compared to placebo, with improved quality of life and fewer adverse events. |
| 16 | Abreu et. Al. 2019 a | USA | Crohns Disease | Induction | 191 | 93 | 98 | Mirikizumab | 127 | Placebo | 64 | High | *Mirikizumab* significantly improved endoscopic and clinical outcomes in patients with moderate-to-severe Crohn's disease, with durable efficacy through Week 52. |
| 16 | Abreu et. Al. 2019 b | USA | Crohns Disease | Maintainence | 191 | 93 | 98 | Mirikizumab | 127 | Placebo | 64 | High | *Mirikizumab* significantly improved endoscopic and clinical outcomes in patients with moderate-to-severe Crohn's disease, with durable efficacy through Week 52. |
| 17 | Sandborn et. Al. 2020 | Australia, Belgium, Canada, Czech Republic, Denmark, Georgia, Hungary, Japan, Lithuania, Moldova, The Netherlands, Poland, the United Kingdom, and the United States. | Ulcerative Colities | Induction | 249 | 149 | 100 | Mirikizumab | 186 | Placebo | 63 | High | Mirikizumab showed efficacy in inducing clinical response and remission in patients with moderately to severely active ulcerative colitis, though the primary endpoint (clinical remission at week 12) was not significant. |
| 18 | Lee et. Al. 2025 a | USA, Belgium | Crohns Disease | Induction | 778 | 450 | 328 | Mirikizumab | 579 | Placebo | 199 | High | Treatment with Mirikizumab significantly improved health-related quality of life and work productivity in patients with moderately to severely active Crohn’s disease compared to placebo. |
| 18 | Lee et. Al. 2025 b | USA, Belgium | Crohns Disease | Maintainence | 778 | 450 | 328 | Mirikizumab | 579 | Placebo | 199 | High | Treatment with Mirikizumab significantly improved health-related quality of life and work productivity in patients with moderately to severely active Crohn’s disease compared to placebo. |
| 19 | D'Haens et. Al. 2023 a | USA, Japan, Germany, Latvia | Ulcerative Colities | Induction | 1447 | 881 | 556 | Mirikizumab | 1084 | Placebo | 363 | High | Mirikizumab was more effective than placebo in inducing and maintaining clinical remission in patients with moderately to severely active ulcerative colitis. |
| 19 | D'Haens et. Al. 2023 b | USA, Japan, Germany, Latvia | Ulcerative Colities | Maintainence | 1447 | 881 | 556 | Mirikizumab | 1084 | Placebo | 363 | High | Mirikizumab was more effective than placebo in inducing and maintaining clinical remission in patients with moderately to severely active ulcerative colitis. |
| 19 | D'Haens et. Al. 2023 c | USA, Japan, Germany, Latvia | Ulcerative Colities | Maintainence | 686 | 389 | 299 | Mirikizumab | 494 | Placebo | 192 | High | Mirikizumab was more effective than placebo in inducing and maintaining clinical remission in patients with moderately to severely active ulcerative colitis. |
| 20 | D'Haens et. Al. 2022 a | Netherlands, Canada, Belgium, USA, Italy, Japan, UK, Spain, France, China, and Germany. | Crohns Disease | Induction | 937 | 497 | 434 | Risankizumab | 745 | Placebo | 186 | High | Risankizumab demonstrated significant efficacy and safety in treating participants with moderately to severely active Crohn's disease. |
| 20 | D'Haens et. Al. 2022 b | Netherlands, Canada, Belgium, USA, Italy, Japan, UK, Spain, France, China, and Germany. | Crohns Disease | Induction | 618 | 309 | 309 | Risankizumab | 411 | Placebo | 207 | High | The study found that Risankizumab significantly improved the efficacy and safety outcomes in participants with moderately to severely active Crohn's disease who had previously failed biologic treatments, demonstrating its potential as a viable option for this difficult-to-treat population. |
| 21 | Feagan et. Al. 2018 | Canada, Spain, Belgium, Germany, Netherlands, USA, South Korea, UK, Austria. | Crohns Disease | Induction | 121 | 74 | 47 | Risankizumab | 82 | Placebo | 39 | High | *Extended treatment with intravenous and subcutaneous risankizumab improves clinical remission and response rates in patients with moderate to severe Crohn’s disease* |
| 22 | Louis et. Al. 2024a | Belgium | Ulcerative Colities | Induction | 975 | 586 | 391 | Risankizumab | 650 | Placebo | 325 | High | Risankizumab significantly improved clinical remission rates in patients with moderately to severely active ulcerative colitis compared to placebo |
| 22 | Louis et. Al. 2024a | Belgium | Ulcerative Colities | Maintainence | 548 | 313 | 235 | Risankizumab | 365 | Placebo | 183 | High | Risankizumab maintenance therapy (180 mg and 360 mg) was superior to placebo in maintaining clinical remission and achieving key clinical, endoscopic, and histological-endoscopic outcomes in patients with moderately to severely active ulcerative colitis. |
| 23 | Ferrante et. al. 2023 | Belgium, the UK, the USA, Canada, China, Japan, Argentina, Portugal, and Germany. | Crohns Disease | Maintainence | 462 | 231 | 231 | Risankizumab | 298 | Placebo | 164 | High | Risankizumab maintenance therapy sustains clinical and endoscopic improvements in patients with Crohn’s disease who initially responded to induction therapy. |
| 24 | Peyrin-Biroulet et. Al. 2024 | France | Crohns Disease | Maintainence | 520 | 267 | 253 | Risankizumab | 255 | Ustekizumab | 265 | High | Risankizumab was superior to ustekinumab with respect to clinical remission at week 24 and endoscopic remission at week 48 in patients with moderate-to-severe Crohn’s disease. |
| 25 | Feagan et. al. 2016 a | Multicenter | Crohns Disease | Induction | 741 | 317 | 424 | Ustekizumab | 494 | Placebo | 247 | High | Ustekinumab is effective for inducing and maintaining remission in Crohn’s disease patients who are refractory to other treatments |
| 25 | Feagan et. al. 2016 b | Multicenter | Crohns Disease | Maintainence | 741 | 317 | 424 | Ustekizumab | 494 | Placebo | 247 | High | Ustekinumab is effective for inducing and maintaining remission in Crohn’s disease patients who are refractory to other treatments |
| 25 | Feagan et. al. 2016 c | Multicenter | Crohns Disease | Induction | 628 | 293 | 335 | Ustekizumab | 418 | Placebo | 210 | High | Ustekinumab is effective for inducing and maintaining remission in Crohn’s disease patients who are refractory to other treatments |
| 25 | Feagan et. al. 2016 d | Multicenter | Crohns Disease | Maintainence | 628 | 293 | 335 | Ustekizumab | 418 | Placebo | 210 | High | Ustekinumab is effective for inducing and maintaining remission in Crohn’s disease patients who are refractory to other treatments |
| 25 | Feagan et. al. 2016 e | Multicenter | Crohns Disease | Induction | 397 | 173 | 224 | Ustekizumab | 264 | Placebo | 133 | High | Ustekinumab is effective for inducing and maintaining remission in Crohn’s disease patients who are refractory to other treatments |
| 25 | Feagan et. al. 2016 f | Multicenter | Crohns Disease | Maintainence | 397 | 173 | 224 | Ustekizumab | 264 | Placebo | 133 | High | Ustekinumab is effective for inducing and maintaining remission in Crohn’s disease patients who are refractory to other treatments |
| 26 | Danese et. Al. 2022 | Multicenter | Crohns Disease | Induction | 498 | 215 | 283 | Ustekizumab | 219 | Placebo | 221 | High | Timely escalation of ustekinumab therapy did not result in significantly better endoscopic outcomes compared to symptom-driven therapy |
| 27 | Sandborn et. Al. 2008 | USA, Canada | Crohns Disease | Maintainence | 202 | 104 | 98 | Ustekizumab | 52 | Placebo | 51 | High | Ustekinumab induces a clinical response in moderate-to-severe Crohn's disease, especially in patients previously treated with infliximab |
| 28 | Sandborn et. Al. 2012 a | Multicenter | Crohns Disease | Induction | 658 | 281 | 245 | Ustekizumab | 526 | Placebo | 132 | High | Ustekinumab showed increased clinical response in patients with moderate-to-severe Crohn's disease who had failed TNF antagonist treatment, particularly in the maintenance phase. |
| 28 | Sandborn et. Al. 2012 b | Multicenter | Crohns Disease | Maintainence | 658 | 281 | 245 | Ustekizumab | 526 | Placebo | 132 | High | Ustekinumab showed increased clinical response in patients with moderate-to-severe Crohn's disease who had failed TNF antagonist treatment, particularly in the maintenance phase. |
| 29 | Sands et. Al. 2019 | Multicenter | Crohns Disease | Induction | 961 | 582 | 379 | Ustekizumab | 642 | Placebo | 319 | High | **Ustekinumab was significantly more effective than placebo in inducing and maintaining clinical remission in patients with moderate-to-severe ulcerative colitis, with a comparable safety profile.** |
| 29 | Sands et. Al. 2019 | Multicenter | Crohns Disease | Maintainence | 961 | 582 | 379 | Ustekizumab | 642 | Placebo | 319 | High | **Ustekinumab was significantly more effective than placebo in inducing and maintaining clinical remission in patients with moderate-to-severe ulcerative colitis, with a comparable safety profile.** |

Table S2. Complete Remission of Chrons Disease in Induction Treatment

|  | Guselkumab | Mirikizumab | Placebo | Risankizumab | Ustekizumab |
| --- | --- | --- | --- | --- | --- |
| Guselkumab | Guselkumab | 1.69 (0.5, 7.29) | 0.33 (0.17, 0.74) | 0.77 (0.35, 2.08) | 0.54 (0.26, 1.37) |
| Mirikizumab | 0.59 (0.14, 2.01) | Mirikizumab | 0.19 (0.06, 0.57) | 0.46 (0.13, 1.51) | 0.32 (0.1, 1.01) |
| Placebo | 3.07 (1.35, 5.98) | 5.19 (1.75, 16.71) | Placebo | 2.38 (1.42, 3.98) | 1.67 (1.2, 2.39) |
| Risankizumab | 1.29 (0.48, 2.89) | 2.18 (0.66, 7.89) | 0.42 (0.25, 0.7) | Risankizumab | 0.7 (0.38, 1.32) |
| Ustekizumab | 1.84 (0.73, 3.84) | 3.11 (0.99, 10.48) | 0.6 (0.42, 0.84) | 1.43 (0.76, 2.63) | Ustekizumab |

Figure S1. Complete Remission of Chrons Disease in Induction Treatment SUCRA Ranking

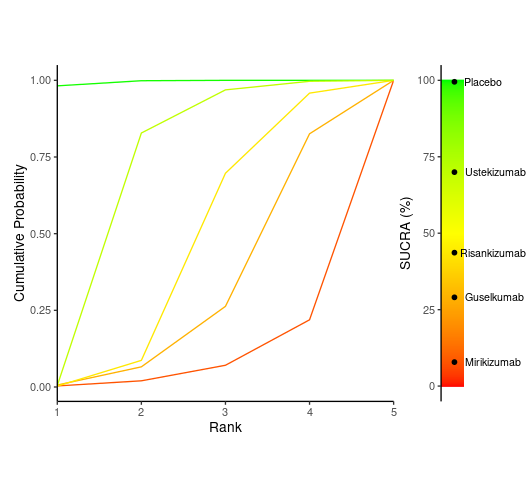

Table S3. Complete Remission of Chrons Disease in Maintainence Treatment

|  | Guselkumab | Mirikizumab | Placebo | Risankizumab | Ustekizumab |
| --- | --- | --- | --- | --- | --- |
| Guselkumab | Guselkumab | 1.69 (0.5, 7.29) | 0.33 (0.17, 0.74) | 0.77 (0.35, 2.08) | 0.54 (0.26, 1.37) |
| Mirikizumab | 0.59 (0.14, 2.01) | Mirikizumab | 0.19 (0.06, 0.57) | 0.46 (0.13, 1.51) | 0.32 (0.1, 1.01) |
| Placebo | 3.07 (1.35, 5.98) | 5.19 (1.75, 16.71) | Placebo | 2.38 (1.42, 3.98) | 1.67 (1.2, 2.39) |
| Risankizumab | 1.29 (0.48, 2.89) | 2.18 (0.66, 7.89) | 0.42 (0.25, 0.7) | Risankizumab | 0.7 (0.38, 1.32) |
| Ustekizumab | 1.84 (0.73, 3.84) | 3.11 (0.99, 10.48) | 0.6 (0.42, 0.84) | 1.43 (0.76, 2.63) | Ustekizumab |

Figure S2. Complete Remission of Chrons Disease in Maintainence Treatment

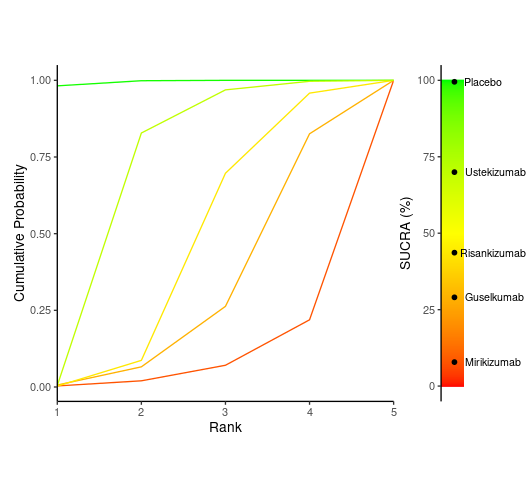

Table S4. Complete Remission of Ulcerative Colitis in induction treatment

|  | Guselkumab | Mirikizumab | Placebo | Risankizumab |
| --- | --- | --- | --- | --- |
| Guselkumab | Guselkumab | 0.39 (0.08, 2.9) | 0.26 (0.08, 0.91) | 1.05 (0.12, 9.11) |
| Mirikizumab | 2.56 (0.35, 12.54) | Mirikizumab | 0.67 (0.15, 2.01) | 2.68 (0.24, 19.34) |
| Placebo | 3.8 (1.1, 13.26) | 1.5 (0.5, 6.63) | Placebo | 3.96 (0.69, 23.03) |
| Risankizumab | 0.96 (0.11, 8.18) | 0.37 (0.05, 4.12) | 0.25 (0.04, 1.46) | Risankizumab |

Figure S3. SUCRA - Complete Remission of Ulcerative Colitis in induction treatment

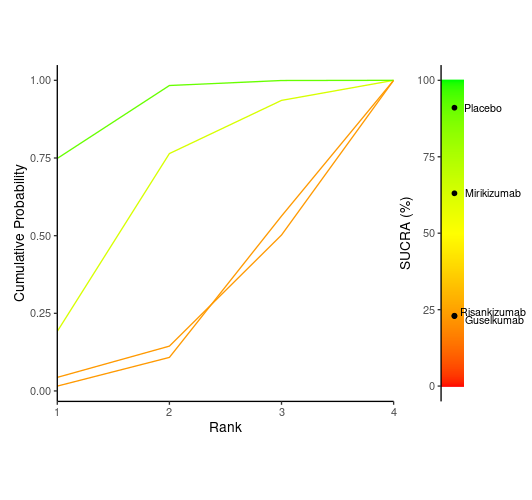

Table S4 Complete Remission of Ulcerative Colitis in Maintainence Treatment

|  | Guselkumab | Mirikizumab | Placebo | Risankizumab |
| --- | --- | --- | --- | --- |
| Guselkumab | Guselkumab | 0.44 (0.07, 2.69) | 0.33 (0.07, 1.43) | 0.62 (0.08, 4.99) |
| Mirikizumab | 2.28 (0.37, 13.73) | Mirikizumab | 0.75 (0.27, 2.05) | 1.43 (0.24, 8.36) |
| Placebo | 3.07 (0.7, 13.38) | 1.34 (0.49, 3.7) | Placebo | 1.9 (0.46, 8.15) |
| Risankizumab | 1.61 (0.2, 12.55) | 0.7 (0.12, 4.19) | 0.53 (0.12, 2.2) | Risankizumab |

Figure S4. SUCRA ranking of Complete Remission of Ulcerative Colitis in Maintainence Treatment

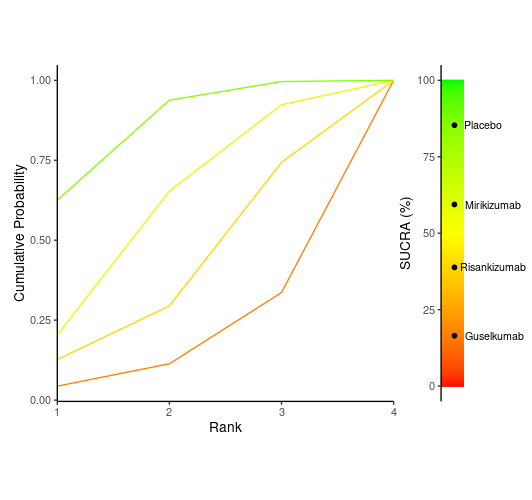

Table S5. Endoscopic Remission of Chrons Disease in Induction Treatment

|  | Guselkumab | Mirikizumab | Placebo | Risankizumab | Ustekizumab |
| --- | --- | --- | --- | --- | --- |
| Guselkumab | Guselkumab | 3.15 (0.39, 77.18) | 0.18 (0.08, 0.36) | 0.59 (0.23, 1.4) | 0.34 (0.14, 0.75) |
| Mirikizumab | 0.32 (0.01, 2.57) | Mirikizumab | 0.06 (0, 0.39) | 0.19 (0.01, 1.39) | 0.11 (0, 0.77) |
| Placebo | 5.68 (2.76, 12.28) | 17.84 (2.57, 404.61) | Placebo | 3.4 (2, 5.44) | 1.92 (1.38, 2.67) |
| Risankizumab | 1.69 (0.71, 4.27) | 5.34 (0.72, 129.82) | 0.29 (0.18, 0.5) | Risankizumab | 0.56 (0.32, 1.06) |
| Ustekizumab | 2.97 (1.33, 6.91) | 9.28 (1.3, 214.57) | 0.52 (0.37, 0.72) | 1.77 (0.94, 3.14) | Ustekizumab |

Figure S5. Endoscopic Remission of Chrons Disease in Induction Treatment SUCRA

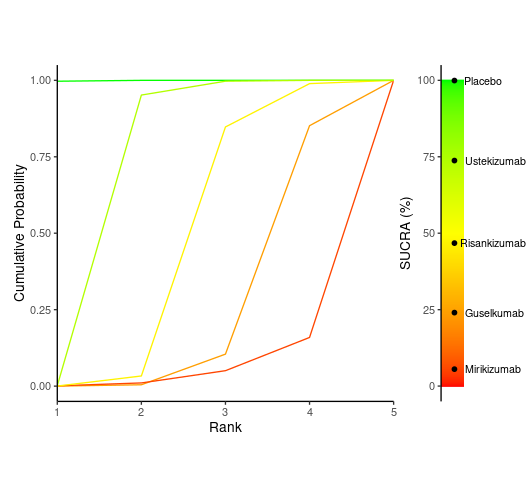

Table S6. Endoscopic Remission of Chrons Disease in Maintainence Treatment

|  | Guselkumab | Mirikizumab | Placebo | Risankizumab | Ustekizumab |
| --- | --- | --- | --- | --- | --- |
| Guselkumab | Guselkumab | 0.04 (0.01, 0.3) | 0.1 (0.02, 0.41) | 0.24 (0.04, 1.31) | 0.12 (0.03, 0.56) |
| Mirikizumab | 24.97 (3.38, 191.07) | Mirikizumab | 2.41 (0.63, 9.46) | 6.02 (1.15, 30.82) | 3.08 (0.71, 13.02) |
| Placebo | 10.3 (2.46, 45.92) | 0.42 (0.11, 1.6) | Placebo | 2.49 (0.95, 6.2) | 1.28 (0.76, 2.09) |
| Risankizumab | 4.15 (0.76, 24.4) | 0.17 (0.03, 0.87) | 0.4 (0.16, 1.05) | Risankizumab | 0.51 (0.2, 1.32) |
| Ustekizumab | 8.08 (1.79, 39.72) | 0.32 (0.08, 1.41) | 0.78 (0.48, 1.32) | 1.95 (0.76, 4.93) | Ustekizumab |

Figure S6. Endoscopic Remission of Chrons Disease in Maintainence Treatment SUCRA

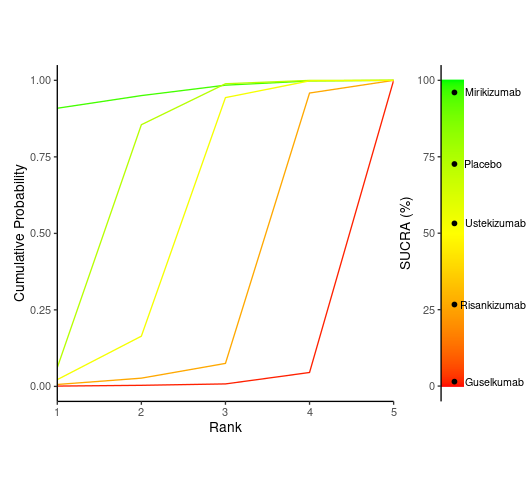

Table S7. Endoscopic Remission of Ulcerative Colitis in Induction Treatment

|  | Guselkumab | Mirikizumab | Placebo | Risankizumab |
| --- | --- | --- | --- | --- |
| Guselkumab | Guselkumab | 0.76 (0.12, 6.1) | 0.3 (0.07, 1.23) | 4.26 (0.38, 48.98) |
| Mirikizumab | 1.31 (0.16, 8.43) | Mirikizumab | 0.4 (0.09, 1.41) | 5.58 (0.45, 57.75) |
| Placebo | 3.32 (0.82, 13.46) | 2.52 (0.71, 11.25) | Placebo | 14.11 (1.99, 102.6) |
| Risankizumab | 0.23 (0.02, 2.64) | 0.18 (0.02, 2.24) | 0.07 (0.01, 0.5) | Risankizumab |

Figure S7. SUCRA - Endoscopic Remission of Ulcerative Colitis in Induction Treatment

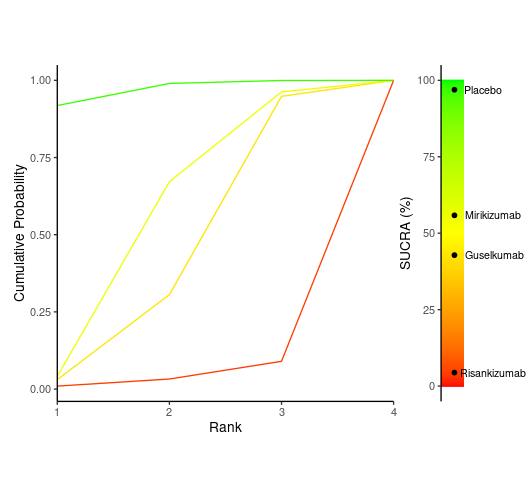

Table S8. Endoscopic Remission of Ulcerative Colitis in Maintaince Treatment

|  | Guselkumab | Mirikizumab | Placebo | Risankizumab |
| --- | --- | --- | --- | --- |
| Guselkumab | Guselkumab | 0.61 (0.14, 2.61) | 0.3 (0.09, 0.99) | 0.64 (0.12, 3.47) |
| Mirikizumab | 1.64 (0.38, 7.08) | Mirikizumab | 0.49 (0.22, 1.1) | 1.06 (0.26, 4.4) |
| Placebo | 3.35 (1.01, 11.29) | 2.04 (0.91, 4.53) | Placebo | 2.16 (0.67, 7.01) |
| Risankizumab | 1.55 (0.29, 8.28) | 0.94 (0.23, 3.9) | 0.46 (0.14, 1.49) | Risankizumab |

Figure S8. SUCRA Endoscopic Remission of Ulcerative Colitis in Maintaince Treatment

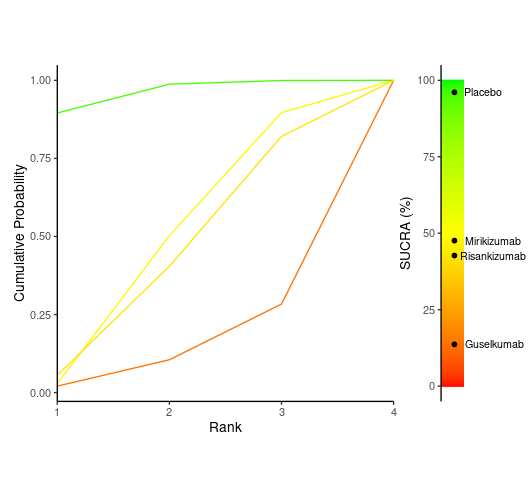

Table S9. Histological Remission of Chrons Disease Maintenance Treatment

|  | Guselkumab | Mirikizumab | Placebo | Risankizumab |
| --- | --- | --- | --- | --- |
| Guselkumab | Guselkumab | 2.24 (0.33, 14.87) | 1.03 (0.26, 3.78) | 2.49 (0.23, 26.09) |
| Mirikizumab | 0.45 (0.07, 3.06) | Mirikizumab | 0.46 (0.12, 1.75) | 1.11 (0.1, 11.82) |
| Placebo | 0.97 (0.26, 3.85) | 2.18 (0.57, 8.48) | Placebo | 2.43 (0.35, 17.22) |
| Risankizumab | 0.4 (0.04, 4.41) | 0.9 (0.08, 9.62) | 0.41 (0.06, 2.88) | Risankizumab |

Figure S9. Histological Remission of Chrons Disease Maintenance Treatment SUCRA

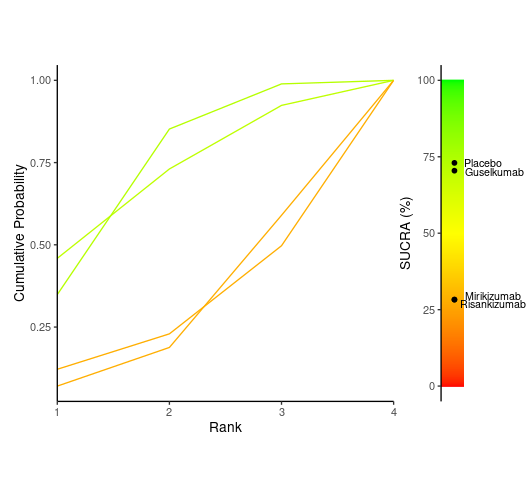

Table S10. Response to Treatment of Chrons Disease for Induction Treatment

|  | Guselkumab | Mirikizumab | Placebo | Risankizumab |
| --- | --- | --- | --- | --- |
| Guselkumab | Guselkumab | 0.75 (0.21, 2.7) | 0.37 (0.13, 1.05) | 0.97 (0.22, 4.24) |
| Mirikizumab | 1.33 (0.37, 4.81) | Mirikizumab | 0.49 (0.24, 1.02) | 1.29 (0.36, 4.6) |
| Placebo | 2.7 (0.95, 7.71) | 2.03 (0.98, 4.18) | Placebo | 2.61 (0.92, 7.39) |
| Risankizumab | 1.03 (0.24, 4.5) | 0.78 (0.22, 2.79) | 0.38 (0.14, 1.09) | Risankizumab |

Figure S10. Response to Treatment of Chrons Disease for Induction Treatment

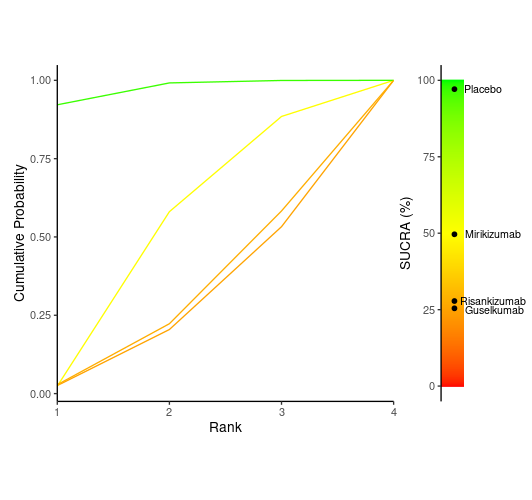

Table S11. Response to Treatment of Chrons Disease for Maintenance Treatment

|  | Guselkumab | Mirikizumab | Placebo | Risankizumab | Ustekizumab |
| --- | --- | --- | --- | --- | --- |
| Guselkumab | Guselkumab | 0.07 (0, 1.43) | 0.19 (0.02, 1.53) | 0.35 (0.02, 6.98) | 0.79 (0.04, 14.68) |
| Mirikizumab | 14.1 (0.7, 268.79) | Mirikizumab | 2.68 (0.31, 23.07) | 4.93 (0.23, 100.67) | 11.1 (0.57, 220.27) |
| Placebo | 5.27 (0.65, 41.8) | 0.37 (0.04, 3.22) | Placebo | 1.82 (0.22, 16.27) | 4.13 (0.54, 32.93) |
| Risankizumab | 2.88 (0.14, 55.8) | 0.2 (0.01, 4.31) | 0.55 (0.06, 4.61) | Risankizumab | 2.26 (0.11, 43.24) |
| Ustekizumab | 1.27 (0.07, 23.85) | 0.09 (0, 1.76) | 0.24 (0.03, 1.87) | 0.44 (0.02, 8.74) | Ustekizumab |

Figure S11. Response to treatment of Chrons Disease for Maintainence Treatment

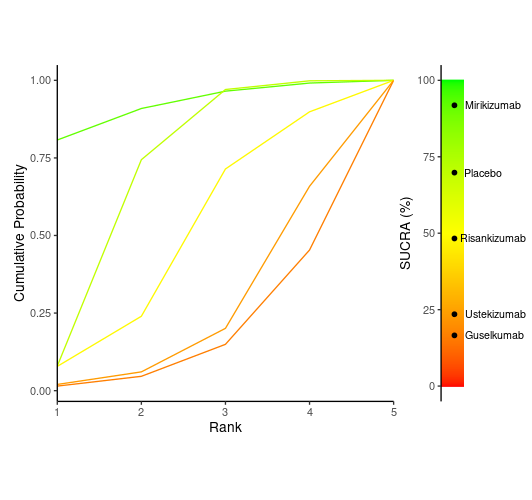

Table S12. Response to Treatment of Ulcerative Colitis for Induction Treatment

|  | Guselkumab | Mirikizumab | Placebo |
| --- | --- | --- | --- |
| Guselkumab | Guselkumab | 0.51 (0.05, 5.43) | 0.24 (0.04, 1.57) |
| Mirikizumab | 1.98 (0.18, 19.14) | Mirikizumab | 0.48 (0.12, 1.73) |
| Placebo | 4.15 (0.64, 27.33) | 2.1 (0.58, 8.65) | Placebo |

Figure S12. SUCRA - Response to Treatment of Ulcerative Colitis for Induction Treatment

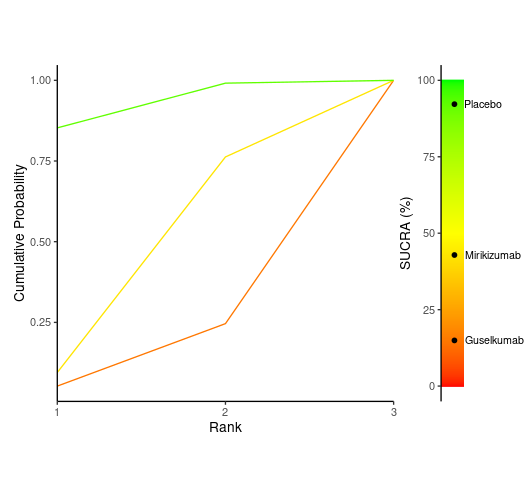

Table S13. Response to Treatment of Ulcerative Colitis for Induction Treatment

|  | Guselkumab | Mirikizumab | Placebo |
| --- | --- | --- | --- |
| Guselkumab | Guselkumab | 0.92 (0.31, 2.79) | 0.58 (0.23, 1.45) |
| Mirikizumab | 1.09 (0.36, 3.23) | Mirikizumab | 0.64 (0.34, 1.15) |
| Placebo | 1.71 (0.69, 4.31) | 1.57 (0.87, 2.94) | Placebo |

Table S13. Response to Treatment of Ulcerative Colitis for Induction Treatment

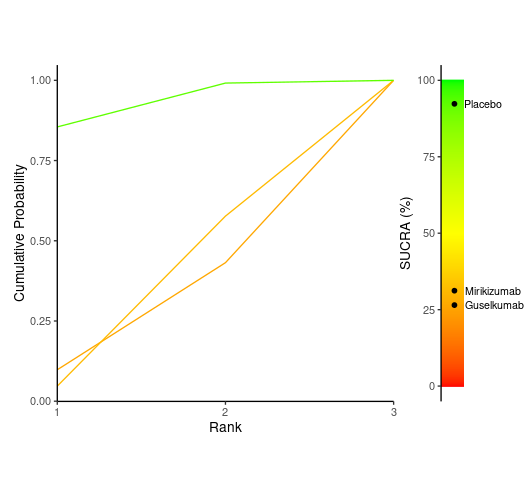

Figure S14. Risk of Bias ROBS 2.0

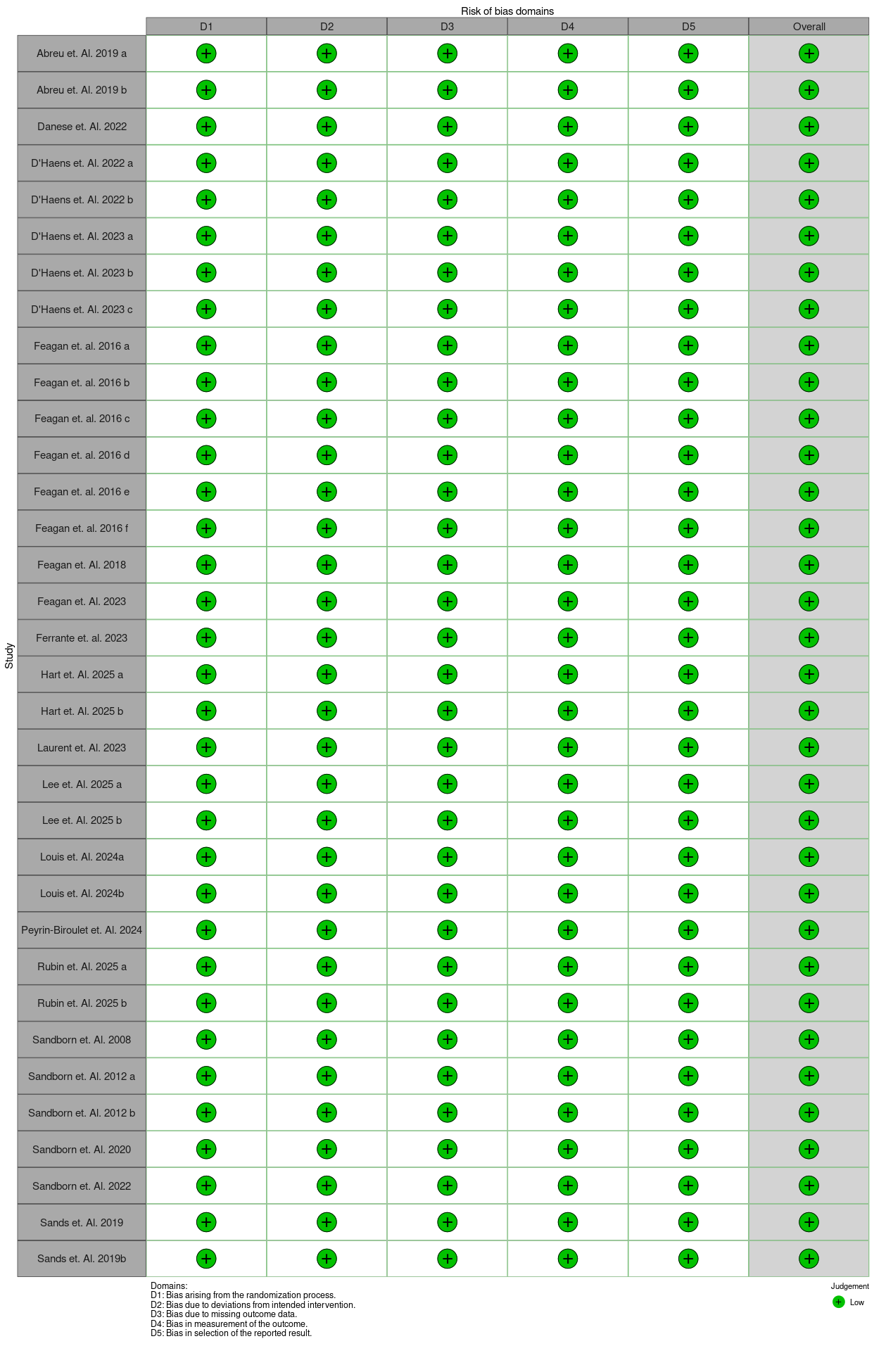
